# A systematic review of the experiences of minority language users in health and social care research

**DOI:** 10.1101/2022.04.27.22273307

**Authors:** Dr Llinos Haf Spencer, Mrs Beryl Ann Cooledge, Dr Zoe Hoare

## Abstract

To obtain the most rigorous results in health and social care research (HSCR), it is important to have a representative sample. Culture can sometimes be overlooked but can be important to the individual concerned. Ethnic minorities are often not accounted for in HSCR. Migration, equality, and diversity issues are now important priorities and therefore need to be considered by researchers. Therefore, it is important to explore the literature examining the experiences of minority language users in HSCR. A systematic review (SR) of the literature was conducted to answer the research question. SPIDER framework and Cochrane principles were utilized for the SR. Five databases were searched, yielding 5311 papers initially. A SR protocol was developed and published in PROSPERO. Available from: https://www.crd.york.ac.uk/prospero/display_record.php?ID=CRD42020225114analysis. Following the title and abstract review by two authors, 74 papers were included, and a narrative account was provided. Six themes were identified: 1. Disparities in health care; 2. Maternal health; 3. Mental health; 4. Methodology in health research; 5. Migrant and minority health care; 6. Racial and ethnic gaps in health care. It was evident that language barriers (including language proficiency) and cultural barriers still exist in terms of recruitment, possibly effecting the validity of the results. Several papers acknowledged language barriers but did not act on this information. Ethnic minorities’ needs are not completely addressed in HSCR. Despite research acknowledging different cultures over the past 40 years, there is a need for this to be fully acknowledged and embedded in the research process. We propose that future research should include details of ethnicity and languages spoken so that readers can understand the sample composition to be able to interpret the results in the best way, recognising the significance of culture and language.

**2-4 Key messages, detailing the main points made in the paper:**

- Ethnic minorities’ needs are not completely addressed in health and social care research. Despite research acknowledging different cultures over the past 40 years, there is a need for this to be fully acknowledged and embedded in the research process.
- Ethnic background and languages spoken by the research participants should be identified and addressed throughout the research process (from design of the study to dissemination of findings).
- Language and cultural preferences should be appropriately considered/included in the analysis.
- Future research should include details of ethnicity and languages spoken so that readers can understand the sample composition to be able to interpret the results in the best way, recognising the significance of culture and language.

## Introduction

### Ethnic minorities in health research overview

Ethnic minorities across the globe encounter disparities in healthcare (Dressler, Oths and Gravlee, 2005; Vandan *et al*., 2020). The completeness of ethnicity data within healthcare and routine databases has historically been poor (Khunti *et al*., 2020). However, the gap between knowledge translation and minority-language speakers is now starting to close with new legislation (Roberts and Burton, 2013) and interest from the research community (Martin, 2021).

For research to improve the health of our communities it needs to serve the interests of all, recognising diversity and acknowledging the importance of culture. Achieving this needs a systematic approach, starting with asking the right questions, designing inclusive trials, removing regulatory, financial, and institutional barriers to inclusion, and most importantly, building long-term relationships with under-served groups. Crucially, we need to listen to what they say. Ensuring that research is designed so that its participants reflect those who might benefit from the results is vitally important (Treweek *et al*., 2021). Treweek et al., (2021) noted that “Thinking about the number of people in our trials is not enough: we need to start thinking more carefully about who our participants are.” (Page 10 of 12 pages).

In 2013, there was a call for action to include ethnic minorities in research (Gill and Redwood, 2013), and by 2021 the framework for including ethnic minorities in health research was developed (Treweek *et al*., 2021). There is increasing evidence to suggest that culture and language-responsive research enhances rigour, inclusivity, and fairness. Thus, engaging with research participants through a language that is meaningful to them is key to good clinical research practice (Irvine *et al*., 2006; Roberts *et al*., 2007). However, there is research from around the world, suggesting that people who speak minority languages are under-represented in health research (Gill and Redwood, 2013). There are varying reasons for this, including issues regarding recruitment, especially when the researcher does not come from the same community as the minority language users and does not share the same culture and contacts.

Pyett (2002) noted that:

> *“It maybe difficult,… for a researcher to establish credibility with a marginalised or minority group unless they are a member of that group. Many conventional research techniques are not appropriate for these groups since language, literacy and cultural difference can lead to misunderstandings and mis-interpretations.” (page 332)* (Pyett, 2002).

The Welsh Government also has policies for an ‘inclusive Wales’ (Welsh Government, 2017) in which all people from all backgrounds, abilities and religions are encouraged to take an active part within in all aspects of community life. The Prosperity for Wales National strategy (Welsh Government, 2017) highlight the following:

> *“Communities prosper where people can participate fully and play an active role in shaping their local environment, influencing the decisions which affect them”. Page 19 of 28*.

Taking part in research is not high on the agenda of the typical member of the public, and seems to be even less so for minority language speakers (Jacobs *et al*., 2006). In Wales, according to the 2011 Census, 19 per cent (562,000) of usual residents in Wales aged three and over reported that they could speak Welsh. Thirty per cent (169,000) of this group were aged between three and 15 years old (Office for National Statistics, 2012). There is currently no evidence to suggest that Welsh speakers do or do not take part in research as the information is not routinely collected as part of the demographic data collection in a health research project or clinical trial. This suggests that cultural sensitivity and language awareness cannot be assumed, and therefore must be measured.

### Theory of Durable Inequality

In his Theory of Durable Inequality (TDI) (Tilly, 1998) Tilly argues that the clumping together of ethnic categories with socio-economic categories helps to reinforce exploitation. This could lead to durable inequalities (Lorant and Bhopal, 2011). The under-serving of ethnic minorities in HSCR is an issue which needs more exploration and explanation especially during this time of increased global mobility and migration (Vandan *et al*., 2020). The aim of this systematic review was to investigate the issues surrounding minority language speakers in HSCR, acknowledging how language and culture makes up the individuals’ identity.

## Materials and methods

A systematic review of the literature was utilised based on the step by step approach published in 2018 (Pati and Lorusso, 2018). Five databases were searched including the following: Applied Social Science Index (ASSIA), CINAHL Plus with Full text, PsycINFO, PubMed and Web of Science (Core Collection). Grey literature was also be sought through the Google Search Engine and reference sections from other papers. The searches were restricted to English or Welsh text, and in terms of time limit, from the beginning of January 2000 to the end of December 2020. A twenty-year time was selected since minority language barriers in research began to be reported around the early 2000’s.

The search strategy was developed in conjunction with a professional librarian at Bangor University, and consists of five levels: population, phenomenon of interest, design, evaluation, and research type, according to the SPIDER framework. The search strategy information is shown in Appendix 1. As we were only interested in patient experience of taking part in studies as minority language users, we are not comparing minority language users with the main linguistic group of their country, therefore there is no control or comparator in this systematic review approach. All the included papers were quality appraised using the appropriate appraisal checklists (See Supplementary Material 1). Ethical approval was not necessary for this SR.

The 74 included studies were a mixture of qualitative studies, quantitative studies, mixed methods studies, and reviews. There were **n=22** qualitative studies; **n=10** quantitative studies (including n=1 cohort study and n=9 survey studies including on-line and pilot surveys), there were **n=5** cross-sectional studies; **n=9** mixed methods studies; **n=1** case report; and **n=27** reviews (including n=7 systematic reviews; n=18 research syntheses and n=2 meta-analyses). See Appendix 2 for included studies by study design.

## Results

The PRISMA (Page *et al*., 2021) diagram is shown in Figure 1 and the list of studies is included in Table 1. An in-depth critical review of the quality of each article was conducted (see Supplementary Material 1) and all relevant information extracted into a Word document according to study design (see Appendix 2). A narrative synthesis of the findings will be presented in this analysis section.

**Figure 1.**
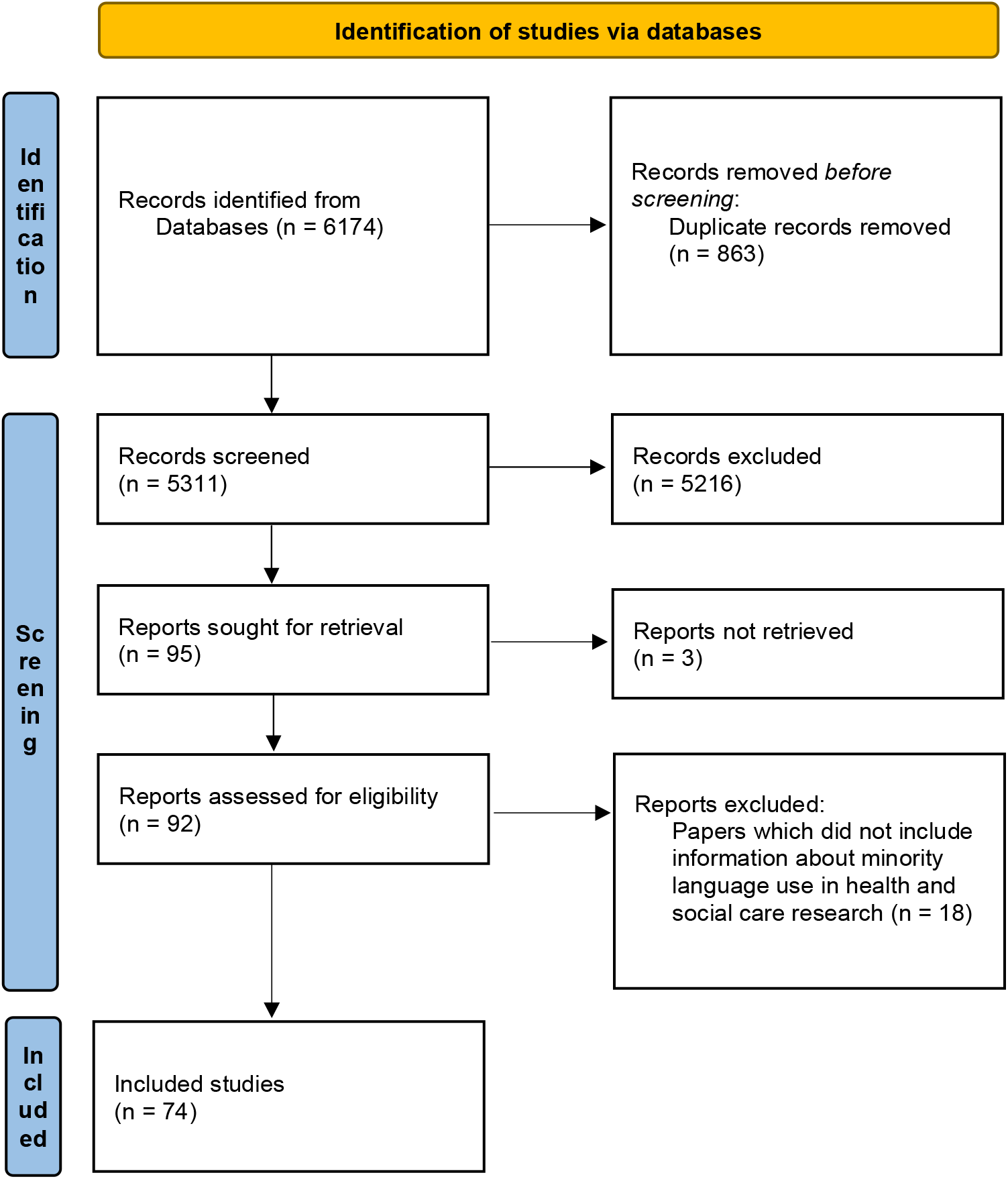
Identification of studies via databases. From: Page MJ, McKenzie JE, Bossuyt PM, Boutron I, Hoffmann TC, Mulrow CD, et al. The PRISMA 2020 statement: an updated guideline for reporting systematic reviews. BMJ 2021;372:n71. doi: 10.1136/bmj.n71 (Page et al., 2021).

For this narrative analysis the 74 included papers were categorised according to the following six themes:

1. Disparities in health care
2. Maternal health
3. Mental health
4. Methodology in health research
5. Migrant and minority health care
6. Racial and ethnic gaps in health care

Further sub-themes were identified within the six main themes (see Table 2).

### Theme 1: Disparities in health care

This theme included two sub-themes including disparities in health care research (n = 8 papers) and disparities in health care use (n = 7 papers). The disparities in health care research papers (Hunt and Bhopal, 2004; Gibbs *et al*., 2006; Jacobs *et al*., 2006; Sue and Dhindsa, 2006; Beresford, 2007; Lau, Chang and Okazaki, 2010; Goode *et al*., 2014; Kaplan, 2014) were all published between 2004 and 2014 and related mainly to the UK and the USA. All these papers (n=7) were research syntheses focussing on the need for the research community to assists health care providers and policymakers with the evidence they require to design and effectively implement linguistically accessible services to limited English proficiency (LEP) patients (Jacobs *et al*., 2006). Even when people belonging to another culture speak fluent English they do not necessarily share the beliefs and values of native English speakers (Hunt and Bhopal, 2004). Authors also highlighted that research in both disability and ethnicity frequently fails to address the multiple cultural identities within population groups (Goode *et al*., 2014).

The disparities in health care use papers (Gaston-Johansson *et al*., 2008; Hahm *et al*., 2008; Coustasse *et al*., 2010; Chirewa, 2012; Akhavan and Karlsen, 2013; Di Pietro and Illes, 2014; Wilk, Maltby and Phillips, 2018) were published between 2008 and 2018. These were a mixture of survey studies, cross-sectional designs, mixed methods studies, qualitative studies, and a systematic review. The systematic review was conducted in 2014 in Canada (Di Pietro and Illes, 2014) and highlighted the paucity of research conducted with Aboriginal groups in Canada. The inclusion of ethnic minority groups should be improved to ensure that the views of minority groups such as the Indian, Inuit and Métis peoples are represented in research planning and decision-making, from the earliest stages of conception and design of projects through to the analysis and dissemination of study results. Survey studies (Hahm *et al*., 2008; Coustasse *et al*., 2010) have also acknowledged that the language used in the survey interviews may affect respondents’ self-reported health status (Coustasse *et al*., 2010). There is also recognition that language spoken is an increasing issue in healthcare research and delivery in the USA due to in-migration from many countries including South American countries, China, Vietnam, Korea and Cambodia (Hahm *et al*., 2008). The qualitative studies (Gaston-Johansson *et al*., 2008; Akhavan and Karlsen, 2013) emphasise the point that the power disparity between doctors and ‘migrant’ patients encourages a sense of powerlessness and mistreatment among patients (Akhavan and Karlsen, 2013). In the USA, authors have also noted that more ethnic minority speakers and bilingual providers should be trained to provide a good health service for minority language speakers (Gaston-Johansson *et al*., 2008). In mixed-methods research, authors have noted that recognizing, respecting, and embracing the different cultures of the stakeholders and partner National Government Organisations (NGOs) is imperative for successful participatory action research Addressing diversity can foster inclusiveness and active participation of members. It is therefore important to reach out to as many NGO forum members as possible (Chirewa, 2012).

### Theme 2: Maternal health

This maternal health theme included three papers which specifically investigated maternal health in relation to minority populations (Strohschein *et al*., 2010; Huang *et al*., 2019; Johnsen *et al*., 2020). One of the papers was a systematic review published in 2019 and two papers were qualitative studies published in 2010 (Strohschein *et al*., 2010) and 2020 (Johnsen *et al*., 2020). The systematic review was a review of 10 qualitative papers which focussed on the language barriers or differences in socio-economic status, and clinical practices that conflicted with local fears and traditional customs. Despite acknowledging different cultures/ethnicity eight studies out of 10 did not report the preferred language of the individuals (Huang *et al*., 2019). One of the qualitative studies was a linguistic validation of different language versions of a questionnaire about reproductive health used in Canada (Strohschein *et al*., 2010). A reproductive health questionnaire was linguistically validated into several different languages and tested with persons monolingual in Hindi, Tamil, Urdu, Spanish, and French in interviews concerning the meaning, clarity, and relevance of instruments. Changes were made to the measures in accordance with the feedback received. The authors highlight the need to be culturally as well as linguistically aware and noted that the research process is influenced by cultural background. In the qualitative study conducted in 2020 (Johnsen *et al*., 2020), eight mini-group interviews with midwives (n = 18) to explore the feasibility of the MAMAACT intervention. This involved a training course for midwives, a leaflet, and a mobile application, as well as additional visit time. This MAMAACT intervention, was developed and tested at a maternity ward to increase responses to pregnancy warning signs among midwives and non-Western immigrant women in Denmark. Three main categories were identified in the qualitative work, these were ‘Challenges of working with non-Western immigrant women’, ‘Attitudes towards and use of the leaflet and mobile application’, and ‘Organisational factors affecting the use of the MAMAACT intervention’. The authors noted that language proficiency was of great importance for the provision of care. Midwives had concerns about communication difficulties causing adverse events. Many non-Western immigrant women were described as lacking the ability to express themselves in Danish or English. Even though the hospital offered interpreter assistance, interpreters were not always available for the midwifery visits. Sometimes, immigrant women would bring their partner, a relative or a friend to interpret for them. This was described as potentially problematic due to the lack of confidentiality and the lack of ability to assess the quality of the translation. Midwives were uncertain if women’s symptoms were described accurately and if their information and advice were conveyed as intended. In situations where no interpreters or family members were available to interpret, midwives would try to get by using gestures or simple words to assess the health of the mother and the baby (Johnsen *et al*., 2020). This example clearly demonstrates the significant importance of language in health care.

### Theme 3: Mental health

The mental health theme included n = 10 papers which specifically investigated mental health in relation to minority populations (Sadavoy, Meier and Ong, 2004; Shattell *et al*., 2008; De La Torre, 2009; Woodall *et al*., 2010; Kim *et al*., 2011; Rose and Cheung, 2012; Brisset *et al*., 2014; Brown *et al*., 2014; Lu *et al*., 2014; Durbin, Sirotich and Durbin, 2017). Two of the papers were systematic reviews (Woodall *et al*., 2010; Brown *et al*., 2014); one paper was a research synthesis (Rose and Cheung, 2012); one paper was a cross-sectional study (Durbin, Sirotich and Durbin, 2017); three of the papers were qualitative studies (Sadavoy, Meier and Ong, 2004; Shattell *et al*., 2008; De La Torre, 2009); and three of the papers used survey methodology (Kim *et al*., 2011; Brisset *et al*., 2014; Lu *et al*., 2014).

The systematic review published in 2010 investigating barriers to taking part in mental health research with particular reference to gender, age, and ethnicity (Woodall *et al*., 2010) and included 49 papers. These papers provided evidence of a wide range of barriers including transportation difficulties, distrust and suspicion of researchers, and the stigma attached to mental illness. Strategies to overcome these barriers included the use of bilingual staff to recruit into mental health research, assistance with travel, avoiding the use of stigmatising language in marketing material and a focus on education about the disorder under investigation. In the SR published in 2014 (Brown *et al*., 2014). The authors noted that it is important that these barriers are considered when planning the research design so that solutions to overcome such obstacles can be incorporated in protocols from the start and appropriate resources allocated. Five of the 9 papers covering Latinos, African Americans and Asian Americans highlighted language related barriers to recruitment. The authors of one paper describing multisite recruitment of Latinos commented, “the issue of communication between the provider/care network staff and the patient with serious mental illness was seen as a potentially overwhelming barrier in many clinical settings” and participants were often monolingual in Spanish or preferred to speak in Spanish. A paper describing recruiting from low-income Latinos, highlighted non-availability of culturally adapted questionnaires as a barrier. A research synthesis published in 2012 (Rose and Cheung, 2012) analysed 54 articles published between 2001 and 2011, and the main focus was on the Diagnostic and Statistical Manual 5 (DSM-5) mental health classification system. Many articles used in this review discussed the inadequacies of the DSM usage, which limited the diagnoses of people of colour and further increased mistrust and disparities. If a person is unable to receive a proper diagnosis of a mental health condition, it is assumed that proper treatment will never be reached. The authors noted that language barriers sometimes stand in the way of elderly people who are not first language English in seeking a diagnosis of a mental health condition (Rose and Cheung, 2012).

A cross-sectional study published in Canada (Durbin, Sirotich and Durbin, 2017) used anonymized routinely collected clinical data from mental health case management programs based in seven community organizations operating in a major metropolitan area in Ontario, Canada. In the bivariate analysis, LEP persons were more likely than English speakers to have unmet needs across all nine need domains (all p values < 0.02) of housing food, transportation and community living skills. Research has shown that across jurisdictions LEP clients with language concordant providers have better outcomes (Durbin, Sirotich and Durbin, 2017).

The three qualitative studies investigating mental health (Sadavoy, Meier and Ong, 2004; Shattell *et al*., 2008; De La Torre, 2009) were conducted between 2004 and 2008 in Canada and the USA. A qualitative action research study was conducted in 2004 in Canada (Sadavoy, Meier and Ong, 2004) using focus group methodology; 17 focus groups were conducted and the authors noted that the dearth of appropriate psychiatrists with language and cultural competency is the most clearly identified gap in mental health service provision. A qualitative study from the USA (Shattell *et al*., 2008) also used focus group methodology to investigate the mental health service needs of a Latino population in 2006-2007. Mental health service needs were discussed by members of the community research team and appropriate solutions were proposed (Shattell *et al*., 2008). Their socio-ecological model was described (P.354), and refers to solutions at the individual, organisation, and community levels. Individual level example included language: Inability to speak English was seen as a barrier to job attainment and ability to access health education messages and mental health services (P.358). Even when an American provider spoke Spanish, the Latinos were apprehensive about sharing their health concerns and viewed them suspiciously.

> *“I meet with somebody; they’re always kind of guarded because they’re wondering ‘What is this American doing in my house? And how is it they speak Spanish? And what’s going on?”’ (P.358)*.

Organisational level example: There are only a few bilingual (healthcare) providers, therefore it is hard to refer. “If you get a Spanish-speaking person in, you are really limited as to who you can refer them to.” (P. 361). Community level example: One of the greatest assets for the Latino community is its own people (Shattell *et al*., 2008). Another qualitative study published in 2009 (De La Torre, 2009) also investigated the concern and needs of Hispanic patients having psychiatric outpatient treatment. The investigators were interested in medication compliance, psychiatric treatment, and recovery from severe mental health disorder. They found that language barriers constituted an obstacle to treatment progress because it triggered poor relationships with the psychiatrist, poor quality of care, participants’ inability to appropriately express their feelings and needs, poor communication between participants and their psychiatrist, and at times, participants’ early termination of treatment. Quotes included:

> *“She does not speak Spanish. I would like someone that I could share my problems and feelings in my language. It is hard for the doctor to understand what I am saying due to my accent…. I would like to have a real conversation. I would like to speak with someone that is really interested in my problems, fears, and anxieties.” (P.234)* (De La Torre, 2009)

Also, one female respondent complained about the lack of rapport that she had with her psychiatrists. She did not speak Spanish; thus, the respondent needed a translator in each appointment. The respondent claimed that the psychiatrist *“was not sensitive”* (P.172) to her limited English skills, and sometimes made her *“feel uncomfortable.” (P.259)* (De La Torre, 2009).

Three surveys also investigated mental health and minority languages (Kim *et al*., 2011; Brisset *et al*., 2014; Lu *et al*., 2014) between 2011 and 2014. The 2011 survey (Kim *et al*., 2011) examined the effect of LEP on mental health service use among immigrant adults with psychiatric disorders. Drawn from the National Latino and Asian American Study (NLAAS), Latino and Asian immigrant adults. English-speaking ability was assessed using a single question “How well do you speak English?” Responses were dichotomized into “excellent/good” or “fair/poor.” Those who reported their English-speaking ability as fair/poor were deemed to have LEP. Mental health was also self-rated. The authors found that LEP was a barrier to mental health service use among Latino immigrants with psychiatric disorders. The Canadian survey conducted in 2014 (Brisset *et al*., 2014) was developed to address barriers in accessing mental health care. Having access to interpreters was considered as the most important resource to overcome language barriers, but the great majority of practitioners had not been trained to work with interpreters. Most interpreted consultations involved ad hoc interpreters drawn from the client’s family members or friends. The authors noted that this finding was consistent with the literature regarding ad hoc interpreters being able to offer immediate availability (being present at the same time as the client), continuity (being present for each consultation), and trusted by clients. Disadvantages being that ad hoc interpreters do not necessarily convey clients’ disagreement or resistance about the diagnostic process or treatment (Brisset *et al*., 2014). Another survey conducted in 2014 also investigated the emotional health, treatment seeking, and barriers to accessing mental health treatment among Chinese-speaking international students in Australia (Lu *et al*., 2014). The authors noted that Chinese-speaking international students are a high-risk group for developing psychological distress, yet they tend to underuse mental health services. Common cultural barriers reported by the respondents include not perceiving symptoms as severe or serious enough to warrant treatment. Language difficulties was also highlighted as a barrier to seeking treatment or support (Lu *et al*., 2014).

### Theme 4: Methodology in health research

This theme included n = 14 papers (Wells and Zebrack, 2007; Murray and Buller, 2007; Martinez, Carter-Pokras and Brown, 2009; Tadić *et al*., 2010; Yildiz and Bartlett, 2011; Dingoyan, Schulz and Mosko, 2012; Nishita and Browne, 2013; Waheed *et al*., 2015; Morville and Erlandsson, 2016; Mosconi *et al*., 2016; Schildmann *et al*., 2016; Squires *et al*., 2019; Mowlabaccus and Jodheea-Jutton, 2020; Premji *et al*., 2020). Of these papers focussing on methodology, n=8 were research syntheses published between 2007 and 2020 (Murray and Buller, 2007; Wells and Zebrack, 2007; Martinez, Carter-Pokras and Brown, 2009; Yildiz and Bartlett, 2011; Nishita and Browne, 2013; Waheed *et al*., 2015; Morville and Erlandsson, 2016; Premji *et al*., 2020). The most recent research synthesis was published in 2020 (Premji *et al*., 2020). This was a review of the authors six own papers on the topic of language barriers in qualitative health research. There were between 25 and 110 participants in each of the included papers. Premji et al., (2020) noted that there remain gaps and debates with respect to the relevant ethical and methodological guidance set forth by funding agencies and institutions and proposed in the scientific literature. In Canada, the Tri-Council Policy Statement or TCPS 2 (2014) on ethical conduct for research involving humans, notes that researchers shall not exclude individuals from the opportunity to participate in research based on attributes such as culture, language, religion, race, disability, sexual orientation, ethnicity, linguistic proficiency, gender, or age, unless there is a valid reason for the exclusion. (article 4.1) (Premji *et al*., 2020). A scoping study published in 2016 (Morville and Erlandsson, 2016) assessed methodological challenges when doing research that includes ethnic minorities. The literature (n=21 studies) came from the American continent, but also from the Netherlands and Sweden. Five themes were identified including the following of relevance to this systematic review: Defining and recruiting samples: Not many papers defined ‘ethnic minorities’ which influences sampling and recruitment into the research studies. Also, very few instruments and assessments have had their psychometric properties tested regarding the use of language and the understanding of the concepts. Using an interpreter during data collection results in some of the same problems as the translation of questionnaires. Using a verbatim translation during an interview will not make sense in most languages; and as with the questionnaires, the interpretation should capture the meaning of the interviewer’s question. However, the problem of using this approach might give the interpreter the responsibility of taking on a leading role, explaining the meaning of concepts and questions, and thus putting several roles upon the interpreter, leaving a large margin for errors. The results showed methodological issues concerning the entire research process from defining and recruiting samples, the conceptual understanding, lack of appropriate instruments, data collection using interpreters to analysing data. In terms of language issues, authors have described the option of using bilingual interviewers from the same ethnic minority group, but as with interpreters, the results are dependent on the interviewers’ style and cultural knowledge of both cultures (P.412) (Morville and Erlandsson, 2016).

Three qualitative studies published between 2012 and 2015 investigated methodological issues (Dingoyan, Schulz and Mosko, 2012; Schildmann *et al*., 2016; Squires *et al*., 2019). In 2012 a qualitative study using focus group methodology to identify potential participation barriers of people with Turkish migration backgrounds living in Germany was conducted. The authors noted that “language barriers are of relevance with regard to recruitment processes and interviews”. (P. S9). The findings on the perception of the focus groups about how health research is perceived by individuals with Turkish migration backgrounds and the implications for successful recruitment offer considerable recommendations for enhancing participation rates in future research. Feelings of mistrust and anxiety were expressed. Research staff need to build trust and include members of the target population. The focus groups with Turkish women were not conducted in the Turkish language, but the authors noted that: “It was also repeatedly stated that the interviewer should be able to speak the Turkish language” (P. S8). (Dingoyan, Schulz and Mosko, 2012).

Another qualitative paper published in 2016 was a bi-national (United Kingdom/Germany) cognitive interview study using ‘think aloud’ and verbal probing techniques (Schildmann *et al*., 2016). A total of 15 German and 10 UK interviews were conducted. The Integrated Palliative care Outcome Scale was presented in in two languages (in this instance German and English only), enhanced face and content validity through testing more than one language at the same time (Schildmann *et al*., 2016).

Another qualitative study based in the USA (Squires *et al*., 2019) was an analysis of existing semi-structured interview data. The data came from n = 35 home health care providers (31 registered nurses, 3 physical therapists, 1 occupational therapist). This study provides a foundation for examining the intersections of how language barriers may affect quality of care in home health care and the relationship to provider workloads. Barriers included the effort of getting in touch with interpreters; Irritation of having to wait for interpreters (patients and nurses); Paying for interpreters (sometimes the service is there depending on demand) sometimes the health insurance covers it, but not in every state). Problems with interpreter service included threat to patient safety due to accuracy of interpretation (Squires *et al*., 2019).

Two mixed methods studies also looked at methodology in health research (Tadić *et al*., 2010; Mowlabaccus and Jodheea-Jutton, 2020). In 2010 a mixed method study was conducted to investigate the quality of life (QOL) of 44 children and adolescents with hereditary retinal disorders using a generic multidimensional paediatric tool for assessing children’s health related quality of life (HRQoL). The overall participation level was below 50%. In both studies, participants from white ethnic and more affluent socioeconomic backgrounds were overrepresented. The poor recruitment rate from children and adolescents from ethnic minority backgrounds may be because the recruitment letters were sent out in English only (Tadić *et al*., 2010). In 2020, another mixed methods study was undertaken to understand participant perception of clinical research in developing countries (Mowlabaccus and Jodheea-Jutton, 2020). The authors carried out a mixed method study consisting of two phases: a qualitative, with thematic approach followed by a quantitative study with cross-sectional design. A validated questionnaire for India and Korea was adapted for use in Mauritius. Most respondents agreed with the value of research while a minority had poor perception related to trust in research companies and conduct of clinical trials.

Respondents who had previously engaged in clinical research had better knowledge and perception compared to those who did not participate in one. They found a generalized vagueness with regards to the concept of clinical trials, and they noted that informed consent should be in the language of the participant. Regarding perception of clinical trials, and literacy level, a better perception score was found to be associated with a high literacy level (p value = 0.02). (Mowlabaccus and Jodheea-Jutton, 2020).

One survey study investigating methodology (Mosconi *et al*., 2016) was also included. This study was a European multi-language initiative to make the general population aware of independent clinical research: the European Communication on Research Awareness Need (ECRAN) project. The authors searched for, and evaluated, relevant existing materials and developed additional materials and tools, making them freely available under a Creative Commons licence. As the language of medical science is English, people who cannot understand English face a barrier to obtaining intelligible information (Mosconi *et al*., 2016).

### Theme 5: Migrant and minorities in health care

This theme included n = 30 papers which was sub-divided into four sub-categories including Migrant people’s health care needs (n=5); Minority participation in health care (n=7); Minority participation in health care research (n=11); Providing health care services to ethnic minority patients (n=7).

The included papers under the sub-theme of ‘Migrant people’s health care needs’ were published between 2009 and 2021 (MacFarlane, Singleton and Green, 2009; Doyle *et al*., 2013; Thomas *et al*., 2014; Betancourt *et al*., 2015; Hunter-Adams and Rother, 2017). Four of the studies employed a qualitative design and one was a systematic review published in 2021 (Chowdhury *et al*., 2021). In this systematic review unmet healthcare needs among migrant populations in Canada was explored and 31 papers were included in the systematic review. Within their included papers, the authors found five categories of unmet needs across different groups of migrants including immigrants, refugees, and temporary migrants. Immigrants and refugees face unique factors that influence the development of unmet needs, such as socio-cultural differences, communication difficulties, and lack of information. Alternatively, temporary migrants have unmet needs due to factors associated with their immigration clauses, such as healthcare coverage being conditional to work permit renewal or precarious living conditions associated with work-related housing. Overall, Chinese immigrants were the most frequently studied ethnic group (8 of 31 studies) (Chowdhury *et al*., 2021). The four qualitative studies, were published between 2009 and 2017 and were conducted in Ireland and England (MacFarlane, Singleton and Green, 2009); the USA (Doyle *et al*., 2013; Betancourt *et al*., 2015); and in South Africa (Hunter-Adams and Rother, 2017). In the Ireland and England comparison study the authors investigated language barriers in health and social care consultations in the community (MacFarlane, Singleton and Green, 2009). Key findings are that the same range of formal and informal responses to language barriers occurs in practice in both England and Ireland, but proportions of knowledge and use of these responses differ. English service providers have more awareness about the use of formal responses than Irish service providers, but uptake of formal responses remains low in both England and Ireland. There is a need for more attention to the implementation of policies for language barriers in both Ireland and England and further research about the normalization processes (MacFarlane, Singleton and Green, 2009). Example comments from service users:

> *CARe Z6 explained that his wife likes him to interpret to give her confidence and support when describing her problems*.

CARe Z3, who takes her daughter to her GP consultations to interpret said that she does not go to see the doctor if the complaint is of a personal nature that she does not want her daughter to be involved in.

In the USA, two qualitative studies were conducted in 2013 and 2015 (Doyle *et al*., 2013; Betancourt *et al*., 2015). In 2013 researchers investigated the health needs of migrant and seasonal farmworkers by interviewing seasonal farmworkers and healthcare providers. Although not the main focus of the investigation, the social service providers who were interviewed, pointed out service gaps in prenatal, vision, and hearing care, and a lack of Spanish-language health care information for the migrant workers in Texas, USA (Doyle *et al*., 2013). Also, around this time in the USA, a qualitative study addressed health disparities among Somali Bantu and Bhutanese child and adolescent refugees in Greater Boston and Springfield, Massachusetts. The most frequently cited problems were related to language barriers (83%), which were overcome with the help of school personnel and parents (Betancourt *et al*., 2015). The final qualitative study within this methodology theme was a study conducted in South Africa in 2017 (Hunter-Adams and Rother, 2017). The authors conducted semi-structured in-depth interviews and focus groups with people living in Cape Town regarding language barriers of cross-border migrants. The authors found that there were challenges in communication because of a lack of common language. For example, it was reported that participants’ perception of language as a vehicle of discrimination was exacerbated by participants’ experiences of discrimination outside the healthcare system. For example, among Zimbabweans, participants interpreted the language barrier as an unnecessary imposition: Nurses could speak English but tended to communicate in IsiXhosa, even though Zimbabweans could not understand this South African language. In low- and middle-income (LMIC) settings health communication tends to have low priority relative to other pressing issues in the health system. There was no effort to link patients with health care providers which shared the same language (or second language) as them, and because of this many patients felt discriminated against (Hunter-Adams and Rother, 2017).

The included papers under the sub-theme of ‘Minority participation in health care’ (Kale and Syed, 2010; Angus *et al*., 2013; Claydon-Platt, Manias and Dunning, 2013; Wang and Kwak, 2015; Castillo *et al*., 2019; Wang *et al*., 2019; Tatari *et al*., 2020) were published between 2010 and 2020. These were a combination of reviews, mixed methods studies, qualitative studies, and a cross-sectional study.

The meta-analysis published in 2013 (Angus *et al*., 2013) analysed the results from 35 relevant qualitative articles to understand the conditions and conceptualizations of women’s inequitable access to health care in Canada. Eleven of the 35 papers referred to linguistic or cultural constraints. Some authors cited examples of Vietnamese and Chinese immigrant new mothers who did not fully benefit from English language programs and services that were not aligned with their cultural backgrounds. Women valued providers who were culturally sensitive to and astute concerning issues of heteronormativity, ableism, or racism, and they appreciated the opportunity to use trained and confidential interpreters or teletype communication in encounters with health care providers (Angus *et al*., 2013). A more recent SR was published in 2019 (Castillo *et al*., 2019) investigating language barriers in healthcare for Latino immigrants living with spina-bifida. Eighteen papers were included in the SR. Twelve (67%) of the 18 papers reported demographic information related to race/ethnicity. Within the 12 papers that reported this demographic information, only seven articles included Hispanics/Latinos. Of these seven, a single study reported 68% of its participants as being Hispanics/Latinos, however, the remaining studies reported a smaller proportion of Hispanics/Latinos as participants. A high proportion of the included papers contained information on ethnicity. The authors noted that quality of life measures were not in the user’s primary language in many cases and that this should be addressed in future research. As a starting place, the authors would encourage the spina-bifida research community to include Hispanics/Latinos in quality of life studies and conduct interviews and questionnaires in their primary language as an estimated 55 million Hispanics/Latinos live in the USA (Castillo *et al*., 2019).

The cross-sectional study in this theme, was conducted in Norway in 2010 (Kale and Syed, 2010) and investigated language barriers and the use of interpreters in the public health services. The n = 453 participants were health care providers from both primary and specialized healthcare facilities in Oslo, and the survey was conducted in 2004–2005. The authors noted that professional language assistance remains underutilized in the health-care sector. Interpreters could be utilized as cultural mediators. A key area for further improvement was noted as the process of raising awareness among health care providers and institutions regarding the legal responsibility they have to ensure the sufficient level of communication with their patients/clients (Kale and Syed, 2010). Two of the studies in this theme of minority participation in health care were qualitative studies (Claydon-Platt, Manias and Dunning, 2013; Tatari *et al*., 2020). One of the studies was conducted in Denmark (Tatari *et al*., 2020) and the other in Australia (Claydon-Platt, Manias and Dunning, 2013). In 2020, researchers investigated the perceptions about cancer and barriers towards cancer screening among ethnic minority women in a deprived area in Denmark. A total of 37 women from ten different non-Western countries participated in the study based on five semi-structured focus groups, two structured group interviews with an interpreter and three individual interviews with culture experts. Cancer was perceived as a deadly disease that could not be treated. Cancer screening was perceived as only relevant if the women had symptoms. Knowledge about cancer screening was fragmented, often due to inadequate Danish language skills and there was a general mistrust in the Danish healthcare system due to perceived low medical competences in Danish doctors. There was, however, a very positive and curious attitude regarding information about the Danish cancer screening programmes and a want for more information. Inadequate language skills in Danish was one of the four themes emerging.

> *“… So, all the material you get you don’t read it because you don’t understand it … If it is only in Danish, no one looks at it”. (P. 6 of 10).* (Tatari *et al*., 2020)

Some communities depend on the spoken word, rather than the written word. The participants reported that the Arab and Somali communities in Denmark are verbal cultures, and some of the respondents said that they get lots of information by contacting others on social media (Tatari *et al*., 2020). The other qualitative study was conducted in Australia in 2013 (Claydon-Platt, Manias and Dunning, 2013) to investigate the barriers and facilitators people with diabetes from a non-English speaking background experience when managing their medications. They interviewed 11 people with diabetes, 10 carers and 10 health professionals and found that language barriers undermined effective medication management by reducing the quantity and quality of information conveyed to people with diabetes. Likewise, poor communication resulted in non-adherence and, consequently, medication related problems. A number of people with diabetes relied on their carers to relay or translate information (Claydon-Platt, Manias and Dunning, 2013). Two mixed methods studies were conducted in Canada in 2013 and 2015 (Wang and Kwak, 2015; Wang *et al*., 2019). In 2015 a mixed-method study was conducted which captured insights into the experience of Korean immigrants in seeking and receiving healthcare in Canada (Wang and Kwak, 2015). Eight focus groups were conducted with 54 participants (81.5% female and 18.5% male). The authors found that almost all the participants preferred to have a Korean-speaking family physician, while six out of 10 were able to find one. Language barriers, such as difficulty describing symptoms in English, lack of familiarity with medical terms, and difficulty understanding physicians’ instructions in English, were experienced by both long term and new immigrants.

> *“I have lived here a long time, so my English is okay for basic things. But when my symptoms are very complicated and complex, it is true that my English is not sufficient. It is not the same as being a Canadian whose mother tongue is English. I cannot express completely my symptoms to the doctors. That is something I have experienced hundreds of times”. (P.344)* (Wang and Kwak, 2015)

There is some provision for South Korean immigrants in Canada (4 out of 10 people could access a Korean speaking general practitioner), however 6 of 10 could not. Therefore, patients with children tended not to access healthcare in Canada. Nearly half of the participants indicated that they did not receive timely care in Canada.

> *The “long wait time” (P.345) for diagnosis, treatment and operation was the main reason for seeking alternative resources in South Korea, where people can “get the service the day they arrive.” (P.345)* (Wang and Kwak, 2015)

The findings from this paper highlights the importance of language concordance and training of healthcare professionals in dealing with patients from ethnic minority backgrounds (Wang and Kwak, 2015). The other mixed method study in this theme was also conducted in Canada (Wang *et al*., 2019) and focussed on breast and colorectal cancer screening barriers among immigrants and refugees. This mixed methods study involved a retrospective chart review of client data and two focus groups with Access Alliance primary care providers (PCPs). The inclusion criteria were active clients aged 50–74 from all three Access Alliance Toronto sites, who were defined as having had at least one clinic encounter between November 1st, 2012 and October 31st, 2015 inclusive. Regarding language barriers, providers described the need for interpreters as being important but time and energy consuming. Additionally, participants noted that mammography forms and self-administered Faecal Occult Blood Test (FOBT) kits are only available in English or French.

> *“We have 30 minutes per appointment typically, but when they’re dealing with seven other issues and language line, interpretation services, you don’t necessarily have that much time to explain in detail the importance of screening” (page 477).* (Wang *et al*., 2019)

This moderate quality mixed method study of recent immigrants and refugees identifies barriers to breast and colorectal cancer screening and supports potential solutions including culturally-congruent peer workers (including language specific), targeted screening workshops, and language specific visual screening aids (Wang *et al*., 2019).

In the ‘Minority participation in health care research’ sub-theme (Robinson and Trochim, 2007; Fisher, 2011; Eriksson-Sjöö *et al*., 2012; Greene *et al*., 2013; Ahlmark *et al*., 2014; Carlini *et al*., 2015; French and Stavropoulou, 2016; Blanchet *et al*., 2017; Falla *et al*., 2017; O’Connor, Adem and Starks, 2018) there was one cohort study, three mixed-methods studies, four qualitative studies, and three surveys ranging in publication dates from 2007 to 2019. The cohort study was published in 2017 and was a quantitative exploratory study to identify barriers to participation as well as recruitment strategies to engage minority parents of young children in health-oriented research in Ottowa, Canada (Blanchet *et al*., 2017) including parent and child dyads (n=259). The authors found that direct contact between participants and research team members (e.g. during community events) as well as referrals by someone they trusted (e.g. a friend, community partners) were the most effective recruiting strategies. Most recruiters were Francophones, and this probably explains why they recruited more Francophone than Anglophone mothers (Blanchet *et al*., 2017). Five of the studies included in this theme were conducted in the USA, three were qualitative papers (Robinson and Trochim, 2007; French and Stavropoulou, 2016; O’Connor, Adem and Starks, 2018) and two used survey methodology (Greene *et al*., 2013; Carlini *et al*., 2015). The qualitative studies each investigated the barriers to ethnic minorities taking part in research. Authors noted that there appears to be a difference in the barriers to participation as defined by community members themselves, and health professionals’ perceptions of these barriers (Robinson and Trochim, 2007) and that strong local research culture is needed to recruit participants into studies (French and Stavropoulou, 2016). Adequate time to discuss research projects was also seen as a way to increase recruitment of minority groups into studies (O’Connor, Adem and Starks, 2018). In a survey study in 2013, authors also noted that having congruence between the researcher and the participant aided satisfaction with the research (Greene *et al*., 2013) and workforce planning – to recruit health care workers and researchers skilled in Spanish and English may be more cost beneficial in Spanish speaking areas than providing translation services. Similarly in 2015, researchers found that advertising in minority languages could boost recruitment into studies (Carlini *et al*., 2015) e.g. Portuguese adverts on Facebook, and Google and e-mail (Carlini *et al*., 2015). Research from The Netherlands (Falla *et al*., 2017) and Sweden (Eriksson-Sjöö *et al*., 2012) supports the stance that patients engage more with the research process if translators or translated materials are provided in different languages for different communities (Falla *et al*., 2017). Other researchers have noted that being able to opt to be interviewed and give verbal consent to participate in a language other than English, facilitated participation by lay community members in Australia (Fisher, 2011). Also, researchers in Denmark suggested that survey response rate could perhaps increase with the use of different language versions of the national health survey (Ahlmark *et al*., 2014)

The ‘Providing health care services to ethnic minority patients’ sub-theme included n=8 studies (Peek *et al*., 2012; Rebecca J. Schwei *et al*., 2016; Du, 2018; Bhuiyan *et al*., 2019; Tan and Denson, 2019; Joo and Liu, 2020; Silveira *et al*., 2020; Vandan *et al*., 2020). All were published between 2012 and 2020. The SR published in 2020 (Joo and Liu, 2020) aimed to identify barriers to providing healthcare services to ethnic minority patients from the perspective of nurses. Eight papers were included in this SR and five common themes were identified in the qualitative studies included: communication issues; unclear, missing, or culturally inappropriate care information and resources; insufficient cultural training and education; challenging therapeutic relationships with patients; and concern about quality of care. Nurses said that it was hard to assess ethnic minority patient’ conditions and the intensity of their pain. This difficulty providing comprehensive assessment negatively influenced nurses’ ability to provide appropriate care services. The SR also highlighted that on-site interpreters were not always available, and telephonic interpreters were unable to translate complex health care issues or to see facial expressions (Joo and Liu, 2020). In a research synthesis from 2016 (Rebecca J. Schwei *et al*., 2016) the authors noted that there is enough evidence of language barriers in health research by now, therefore future research should concentrate on the effectiveness and cost-effectiveness of providing language concordant care (Rebecca J. Schwei *et al*., 2016). Two cross sectional studies from the USA (Peek *et al*., 2012; Silveira *et al*., 2020) also acknowledge that making an effort to include minority groups in research is one way to address racial and ethnic health disparities (Peek *et al*., 2012). Authors of a cross-sectional oral health study and the Hispanic community found that similar to previous studies, Spanish language preference (lower acculturation) was associated with poor health related quality of life (HRQOL) and that culturally specific interventions aimed at improving oral health and preventing adverse consequences were needed (Silveira *et al*., 2020).

Two mixed-methods studies were included in the ‘Providing health care services to ethnic minority patients’ sub-theme (Bhuiyan *et al*., 2019; Tan and Denson, 2019). The mixed-methods study published in 2019 was conducted in Bangladesh to assess whether medical language is a barrier to receiving healthcare services in Bangladesh (Bhuiyan *et al*., 2019). Despite two languages being used in the medical context (English and Bengali) in Bangladesh, there are many minority languages that are not used in the medical context. The qualitative findings to show that at least some respondents would have liked all their medical instructions in Bengali. Medical language is already different from the native language people speak in their day-to-day conversations. As such, the language difference between doctors and patients sometimes becomes a strong barrier to achieving a successful treatment outcome. The authors also noted that there is strong evidence from around the world that has proven that miscommunication among health providers and patients plays a major role in the healthcare system, and that medical language is acting as a barrier to achieving an effective health service (Bhuiyan *et al*., 2019). A mixed-method study investigated the bilingual and multilingual skills of psychologists practicing in Australia (Tan and Denson, 2019). The authors found that most participants trained in English. They expressed concerns about their application of psychological concepts in other languages, despite good conversational fluency. Participants highlighted language barriers to entering the profession; limited multicultural and multilingual training and supervision in Australia; and the need for more transcultural mental health resources, particularly for small/new migrant communities and people outside large cities. The authors stated that the Psychology profession must actively support supervision, professional development, and practice in community languages (Tan and Denson, 2019). A similar finding was noted in the qualitative study from Hong Kong, China, published in 2020 (Vandan *et al*., 2020). The authors noted that cultural competency training and education provision should be provided for those caring for South Asian patients in Hong Kong (Vandan *et al*., 2020). The Du (2018) case report was a recent reflection of a graduate medical student who provided real-life examples regarding language and communication. The author noted that language concordant care providers and professional medical interpreters are invaluable for LEP families. Healthcare teams can also come together to create inclusive environments that address language barriers and beyond for LEP patients. Language barriers are not the only barriers that LEP persons experience in healthcare. Of note, having language-concordant providers is of even greater importance to LEP patients who also have limited education, indicating greater vulnerability to health disparities (Du, 2018).

### Theme 6: Racial and ethnic gaps

This final theme included n=2 papers, with two-sub-themes. One of the sub-themes was Racial and ethnic gaps in health care (n=1); and the other was Racial and ethnic gaps in health research (n=1). One meta-analysis reporting on racial and ethnic gaps in health care was published in 2013 (Clarke *et al*., 2013). Eleven SR’s in this meta-analysis; n=391 articles in total. Clarke et al (2013) used qualitative theme analysis to develop a taxonomy of disparities interventions and categorized the 391 articles accordingly. The taxonomy consisted of three components: the intervention (e.g., communication-skills training), the strategy, or a group of tactics sharing common characteristics (e.g., delivering education and training), and the level, or who or what was targeted by the effort (e.g., the provider). The most common strategy to improve minority health was delivering education and training (37%). The most common intervention was delivering education about a disease (14%), followed by education in disease self-management (11%). Training in communication-skills (3%) and use of decision-making aids (1%) were less frequent education interventions.

A qualitative study from the USA reporting on racial and ethnic gaps in health research was published in 2017 (Haley *et al*., 2017). Two semi-structured focus groups were conducted with a purposive sample of 29 clinical research coordinators (CRCs) at consecutive international stroke conferences in 2013 and 2014 to gain in-depth understanding of coordinator-level barriers to racial-ethnic minority recruitment and retention into neurological trials. Barriers identified related to translation, literacy, family composition, and severity of medical diagnosis. Potential strategies to overcome these barriers included a focus on developing personal relationships with patients, community and patient education, centralized clinical trial administrative systems, and competency focused training and education for CRCs. The CRCs identified numerous recommendations to improve study recruitment including the need for: advanced preparation, patient education, community education, relationship building with patients and hospital staff, as well as improved CRC hiring using recommended competency assessments, and training. Low literacy levels was perceived as the main barrier to recruitment (Haley *et al*., 2017).

## Discussion

The papers included in this SR highlight the fact that language barriers still exist in health care research despite nearly forty years of progress in this area (Clarke *et al*., 2013). Authors in this review have noted that during the past 40 years the focus on disparities in ethnic minority population research has been on the participant, however, future research should also focus on methods of recruitment, recruitment of researchers and workforce planning in health services and systems in order to serve minority patients appropriately (Clarke *et al*., 2013).

Despite some researchers acknowledging different cultures, there is a need for this to be fully addressed and embedded throughout each stage of the research process. The language barrier issue is only one obstacle in getting people from minority backgrounds to take part in research. Other barriers include cultural and research barriers (Squires, 2009; Squires *et al*., 2019), these will be discussed below.

Language barriers between researchers and participants present significant methodological challenges for researchers undertaking cross-language qualitative studies (Squires, 2009; Squires *et al*., 2019). Tatari’s paper on cancer screening (Tatari *et al*., 2020) showed that if the information sheet was not available in the participant’s language, potential participants would disregard the study and opt not to partake. Similarly, Squires (2009) noted that for multi-language studies, the study’s rigor improves if the investigators explain why they chose one language for the analysis in place of another (Squires, 2009). Explaining this choice is critical if the analysis did not take place in the same language as that of the participants. It clarifies if the authors made the decision for functional or other reasons.

During recent times, authors have commented on cultural barriers to research and have tried to include aboriginal peoples in academic health research (Di Pietro and Illes, 2014). The ethnicity of the researcher must be considered as this prejudice could be assumed to be eradicated should the participant be of the same ethnicity as the researcher. Not considering the participants’ cultural background violates their fundamental rights to ensure equitable representations in an already marginalised population (Di Pietro and Illes, 2014). Extra efforts to recruit participants from ethnic minorities may be needed and comes with additional costs to the research, which should be considered within the planning stages (Brown *et al*., 2014).

Partnerships between academic researchers and aboriginal peoples need to be fostered to address health related research questions. Some members of the public still have a distrust and suspicion of researchers and this is especially true in mental health research (Woodall *et al*., 2010). Authors have emphasised the need for grant funding bodies to ensure that planned research addresses the evaluation of recruitment strategies to ensure relevant ethnic groups are adequately represented (Woodall *et al*., 2010).

Explanations for disparities in health care use in Sweden was categorized into those reflecting social/structural conditions and the presence/ absence of power and those using cultural/behavioural explanations (Akhavan and Karlsen, 2013). Language issues associated with the diagnostic instruments used in Swedish health care were described as a factor that may lead to inequalities in health and health care. For example, physicians were aware that the questionnaires used to diagnose patients with psychological illnesses were culturally bound: ‘‘We need better tools with easier questions” (Akhavan and Karlsen, 2013). Other authors have also noted that HRQoL measures commonly used in health care research, such as Patient Reported Outcome Measures (PROMs) and Patient Reported Experience Measures (PREMs) should be available in the minority languages of the population under study (Castillo *et al*., 2019) and these should also be culturally appropriate to the target population (Strohschein *et al*., 2010; Morville and Erlandsson, 2016).

This systematic review presents evidence that therapeutic relationships and trust improves when culture is considered (Vandan *et al*., 2020). Researchers frequently fail to include participants who experience language barriers in their projects, in part, because they lack the knowledge and experience to do so (Premji *et al*., 2020). Brown et al (2014) (Brown *et al*., 2014).Authors highlighted in this paper have noted that there is a need to incorporate facilitators to recruitment by organizing researcher training and resource allocation; so that this becomes a pre-emptive measure to counteract barriers rather than a post-event reflection on what the barriers were (Brown *et al*., 2014).

The majority of data collection methods within social and medical sciences are developed with what some authors, call the WEIRD sample, i.e. people within Western Educated, Industrialized, Rich and Democratic societies (Henrich, Heine and Norenzayan, 2010). This gives way to one of the major concerns when doing research, which is whether the constructs that are being studied have the same meaning and value across cultures or even exist in all cultures (Morville and Erlandsson, 2016). In terms of changes to policy and practice, authors (Brown *et al*., 2014) have noted that there are issues that need to be taken into account, such as matching up the ethnicity of the researcher with the ethnic group under study. Making the sample as representative as possible by including ethnic minorities, providing more staff training and promoting more community involvement to increase recruitment rates. Further efforts to improve the quality of research are needed to be useful for decision-makers (Huang *et al*., 2019)

Researchers conducting studies including ethnic minorities should be cognisant of the customs, values, and beliefs of the target group(s) before designing any project. Issues of cross language data collection should be seen as a challenge and not as an obstacle, a stimulus to innovative thought and the development of new techniques of investigation. Cultural and linguistic differences have yet to be incorporated as fundamental to sound public health, primary and secondary care, and health promotion (Hunt and Bhopal, 2004). Individuals’ reactions to illness and discomfort, their concepts of health, their help seeking behaviour is intimately bound up with cultural beliefs, values, and experience (Hunt and Bhopal, 2004).

Despite the historical lack of drive to close the knowledge translation gap between research and minority-language communities within health care systems, durable inequalities still persist in health care around the world, including in Wales, where Welsh is a minority language spoken by 19% of the population (Statistics for Wales, 2012). Health care organisations in Wales now have a statutory duty to deliver equitable Welsh language services (Roberts and Burton, 2013). However, this SR did not yield any relevant papers from Wales, indicating that there is still some way to go in Wales to bring the importance of the Welsh language into focus. In Wales, as in Canada (Office of the Commissioner of Official Languages, 2019), there has been a political momentum to deliver the ‘active offer’ in health care situations. In Canada, the active offer is relevant to the minority languages of English or French (dependent on region), and in Wales, the active offer relates to the use of Welsh as a minority language within a majority English speaking community. The active offer of Welsh is promoted as a policy by the Welsh Government (Welsh Government, 2016) and refers to the act of offering services in Welsh before a patient or client has to ask for it.

The evidence presented in this paper suggests that there are still barriers to minority group representation in HSCR. There is general agreement that all the barriers have been recorded over the past few years and that now, the focus moving forward should be on increased effort to recruit minority language speakers, and record ethnicity in research participation to ensure that research findings are as transparent and robust as possible. We propose that future research should include details of ethnicity and languages spoken. Highlighting this may enable researchers to consider language and culture whilst interpreting the results and formulating the recommendations. Thereby ensuring that language and culture is considered throughout the whole research process.

Based on these results, we recommend that:

1. Ethnic background and languages spoken by the research participants should be identified and addressed throughout the research process (from design of the study to dissemination of findings).
2. Language and cultural preferences are appropriately considered/included in the analysis.

Adopting the same philosophy as the Tri-Council Policy Statement or TCPS 2 (2014) (Premji *et al*., 2020), the authors of this paper propose the use of an acronym as an aide memoir to help researchers to remember about including minority language speakers in HSCR, using the word

## Supporting information

Appendix 1 - Search strategy

Appendix 2 - Study design

## Data Availability

The data underlying this article will be shared on reasonable request to the corresponding author.

## Author contributions

All authors are staff at the North Wales Organisation for Randomised Trials in Health and Social Care (NWORTH), Bangor University, North Wales, UK, and although no authors are from lower-middle-income (LMIC) countries, LMIC countries were included in the Systematic Review. Conception or design of the work: LHS and BAC. Data collection: LHS and BAC. Data analysis and interpretation: LHS, BAC and ZH. Drafting the article: LHS, BAC and ZH. Critical revision of the article: LHS, BAC and ZH. Final approval of the version to be submitted: LHS, BAC and ZH.

## Ethical approval

Ethical approval is not required by our institute for carrying out Systematic Reviews of the literature.

## Funding

This work was funded through the North Wales Trials Unit (NWORTH) funded by Health and Care Research Wales.

## Data Availability

The data underlying this article will be shared on reasonable request to the corresponding author.

## Acknowledgement

Professor Paul Brocklehurst (former Director of the North Wales Trials Unit (NWORTH) for his support during this process.

## Conflict of interest

The authors report no conflict of interest.

### RESEARCH

R – **Respond** to the needs of minority language groups

E – **Educate** the research community to remember about including minority language groups to ensure academic rigour

S – **Reassure** minority language communities that it is **Safe** to take part in research in HSCR E – **Ensure equality**

A – **Aim** to include minority language groups

R – **Representative number** of minority language groups C – **Consider and include minority language groups**

H – **Hear the voices** of the representatives of the minority language groups

**Data availability:** The data underlying this article will be shared on reasonable request to the corresponding author.

## List of appendices

Appendix 1 – Search strategy

Appendix 2 – Included studies by study design

## Notes

### Competing Interest Statement

The authors have declared no competing interest.

