## Appendix 1 - Search strategy for "A systematic review of the experiences of minority language users in health and social care research"

The keyword search strategy was developed in conjunction with a professional librarian at Bangor University, and consists of five levels: population, phenomenon of interest, design, evaluation and research type, according to the SPIDER framework:

**Population**

"African American English" OR "African American language" OR "African American Vernacular” OR “Aboriginal language*” OR “black English” OR “Cornish language” OR “Creole language” OR “Jamaican patois” OR “minority language*” OR “second language*” OR AAE OR Adults OR bilingual* OR Breton OR canad* OR catalan* OR Catalonia OR Catalunya OR Code OR code-switch* OR Cornish OR Creole OR ebonics OR ethnic* OR ethno* OR Gaelic OR hispanic* OR irish OR language* OR Manx OR Occitan OR participa* OR patient* OR patois OR Scottish OR Spanish OR Speak* OR Switch* OR Translanguage* OR use* OR volunteer* OR wales OR Welsh*

**AND**

**Phenomenon of Interest and Design**

“active offer” OR awareness OR BAME OR BME OR Cohort OR control* OR culture* OR dementia OR “ethnic minorities” OR experience* OR health* OR involve* OR literac* OR literate OR offer* OR perception* OR prescribing OR Random OR Social N4 disparities OR Social N4 inequalities OR Social N4 Research OR Co-product* OR Partner* Prudent OR Rigour* OR Together* OR transparen* OR valid* OR wellbeing OR well-being

**AND**

**Evaluation**

barrier* OR benefit* OR care* OR challenge* OR competenc* OR dispar* OR disregard* OR Equal* OR Exclu* OR facilitat* OR harm* OR impruden* OR inattention OR inclusiveness OR inclusivity OR inequalit* OR inferior OR injustic* OR insignifican* OR integrate* OR naiveté* OR need* OR neglect* OR omission* OR opportunit* OR oversight* OR quality OR respect* OR thoughtless* OR value*

**AND**

**Research Type**

“Focus group” OR “health research” OR “Mixed methods” OR Controlled OR health n4 research OR health NEAR/4 research OR Interview OR Mixed-methods OR Project* OR Qualitative OR Quantitative OR Randomised OR RCT OR research* OR study OR survey OR trial* The keywords will be applied to the title and to the abstract, and will be used in combination with MeSH terms.
