## Appendix 2 - Study design for "A systematic review of the experiences of minority language users in health and social care research"

### Appendix 2 Included studies by study design

Tables A1 – A9 are included studies categorised by study design.

### Table A1 Cross-sectional studies (n=5)

| **Citation** | **Country** | **Study details** | **Participants** | **Outcomes** | **Results** | **Overview** |
| --- | --- | --- | --- | --- | --- | --- |
| Durbin, A., Sirotich, F., & Durbin, J. (2017). English language abilities and unmet needs in community mental health services: A cross-sectional study. *The journal of behavioral health services & research*, *44*(3), 483-497.  (Durbin, Sirotich and Durbin, 2017) | Canada | **Type of study:** Cross-sectional study  **Description:** This cross-sectional study used anonymized routinely collected clinical data from mental health case management programs based in 7 community organizations operating in a major metropolitan area in Ontario, Canada. | N = 1449 mental health services clients | Camberwell Assessment of Need, a standardized set of items used to measure client needs for care and support across need domains.  Outcomes included Camberwell need ratings in nine domains, grouped into three broad clusters: basic needs (i.e., securing housing, food, and transportation, community living skill needs (i.e., caring for the self and home, day time activities), and health needs (i.e., managing physical health symptoms, psychological distress, or psychotic symptoms. Clients rated their need as: no need, met need (due to help given), or unmet need/serious problem. | Among the sample of 1449 clients, 11.6% reported a preferred language other than English and were classified as limited English proficiency.  In the bivariate analysis limited English proficiency persons were more likely than English speakers to have unmet need across all nine need domains (all p values < 0.02). | In this cross-sectional study, the statistical analysis showed that those with limited English proficiency were more likely to have unmet health and social care needs.  Research has shown that across jurisdictions limited English proficiency clients with language concordant providers have better outcomes. This research supports that finding. |
| Kale, E., & Syed, H. R. (2010). Language barriers and the use of interpreters in the public health services. A questionnaire-based survey. *Patient education and counseling*, *81*(2), 187-191.  (Kale and Syed, 2010) | Norway | **Type of study:** A cross-sectional study.  **Description:** A quantitative cross-sectional design using a structured questionnaire. | N = 453 participants from both primary and specialized healthcare facilities  The participants were health-care providers in Oslo, and the survey was conducted in 2004–2005 | No health outcomes were investigated | The use of interpreter services seems to be sporadic and dependent on the individual health-care practitioner’s own initiative and knowledge. Many survey participants expressed dissatisfaction with both their own methods of working with interpreters and with the interpreter’s qualifications. | The authors note that professional language assistance remains underutilized in the health-care sector.  Interpreters could be utilized as cultural mediators.  A key area for further improvement is the process of raising awareness among health-care providers and institutions regarding the legal responsibility they have to ensure the sufficient level of communication with their patients/clients. |
| Peek, M. E., Wilson, M. S. C., Bussey-Jones, J., Lypson, M., Cordasco, K., Jacobs, E. A., ... & Brown, A. F. (2012). A study of national physician organizations’ efforts to reduce racial and ethnic health disparities in the United States. *Academic medicine: journal of the Association of American Medical Colleges*, *87*(6), 694.  (Peek *et al.*, 2012) | USA | **Type of study:**  Cross-sectional study  **Description:** To characterize national physician organizations’ efforts to reduce health disparities and identify organizational characteristics associated with such efforts. | N = 167 physician organisations (of differing levels of membership) | No health outcomes were investigated | Overall, the most common themes of organizational activities included health care access and the general topic of health disparities. Communication skills, language barriers, cultural competence, and workforce diversity were frequent themes. Despite the recent attention to racial and ethnic discrimination in health care as a contributor to health disparities,1,24– 27 few organizations identified addressing health care discrimination as a means of reducing health disparities. T | Language was one of the thematic domains in the Taxonomy of Organizational Activities and Thematic Domains of U.S. National Physician Organizations’ Efforts to Address Racial and Ethnic Health Disparities |
| Silveira, M. L., Dye, B. A., Iafolla, T. J., Adesanya, M. R., Boroumand, S., Youngblood, M. E., ... & Garcia, A. I. (2020). Cultural factors and oral health-related quality of life among dentate adults: Hispanic community health study/study of Latinos. *Ethnicity & health*, *25*(3), 420-435.  (Silveira *et al.*, 2020) | USA | **Type of study:** Cross-sectional study  **Description:** cross-sectional analysis of baseline data (2008–2011) from the Hispanic Community Health Study/Study of Latinos (HCHS/SOL), a multicenter, population-based, prospective cohort study of 16,415 Hispanic/Latino adults in the U.S. | N =-16,415 Hispanic/Latino adults in the U.S. | Participants were asked four Oral Health Related QOL (ORHQOL) questions derived from the OHIP (Slade and Spencer 1994). | Overall, 57% of individuals experienced poor OHRQOL in at least one of the domains examined. In multivariable analyses, some elements of higher acculturation were associated with greater food restriction and difficulty doing usual jobs/attending school, but not associated with pain or difficulty chewing, tasting, or swallowing. | Previous studies have found that Spanish language preference (lower acculturation) was associated with poor OHRQOL (Riley et al. 2008) and this study support this finding.  Findings from this study can inform targeted strategies such as culturally-competent dental workforce, community based oral health promotion programs, and patient-centred healthcare approaches to improve OHRQOL in the Hispanic minority group. |
| Wilk, P., Maltby, A., & Phillips, J. (2018). Unmet healthcare needs among indigenous peoples in Canada: findings from the 2006 and 2012 Aboriginal Peoples Surveys. *Journal of Public Health*, *26*(4), 475-483.  (Wilk, Maltby and Phillips, 2018) | Canada | **Type of study:**  Cross-sectional survey design  **Description:** Cross-sectional design using the 2006 and 2012 Aboriginal Peoples Surveys (Statistics Canada 2007; Statistics Canada 2012). | From the 2006 Aboriginal Peoples Surveys (APS) survey, 20,720 respondents were included and 24,150 from the 2012 APS. Overall, in 2006, 11.65% (CI: 11.04, 12.26) of indigenous people in Canada reported UHN in the past 12 months with the proportion of people reporting UHN rising to 13.74% (CI: 12.88, 14.60) in 2012. The increase in the percentage of those reporting UHN from 2006 to 2012 (2.09%) was statistically significant. | No outcome measures – but there was a question on unmet healthcare needs. | The paper reported less than 10 responses regarding language problems (Results not available as the sample was under 10). (Page.480) | In this paper, language was not highlighted as a barrier to healthcare. The most common reasons for unmet health needs were related to availability, and the majority of respondents reported needing care for physical health problems |

### Table A2 Systematic Review papers (n=7)

| **Citation** | **Country** | **Study details** | **Participants** | **Outcomes** | **Results** | **Overview** |
| --- | --- | --- | --- | --- | --- | --- |
| Brown, G., Marshall, M., Bower, P., Woodham, A., & Waheed, W. (2014). Barriers to recruiting ethnic minorities to mental health research: a systematic review. *International journal of methods in psychiatric research*, *23*(1), 36-48.  (Brown *et al.*, 2014) | UK | **Type of study:** Systematic review | 9 papers included in the systematic review | A systematic review to identify barriers in recruiting minority groups into research. No outcome measures used. | The barriers that were identified are not all unique to participants from ethnic minorities, although the way in which they manifest themselves is often distinct in minority groups. It is important that these barriers are considered when designing research design so that solutions to overcome such obstacles can be incorporated in research protocols from the start and appropriate resources allocated.  Five of the 9 papers covering Latinos, African Americans and Asian Americans highlighted language related barriers to recruitment. The authors of one paper describing multisite recruitment of Latinos commented, “the issue of communication between the provider/care network staff and the patient with serious mental illness was seen as a potentially overwhelming barrier in many clinical settings” and participants were often monolingual in Spanish or preferred to speak in Spanish. A paper describing recruiting from low income Latinos, highlighted non-availability of culturally adapted questionnaires as a barrier. A further study, in Asian Americans commented that, Asian American immigrants, especially elders, also have relatively lower literacy levels. | All papers included in the systematic review were from the USA.  It is important to allocate resources so that people from ethnic minority backgrounds can participate in research. |
| Castillo, J., Gandy, K., Bradko, V., & Castillo, H. (2019). Language and Latino immigrants living with spina bifida: Social determinants of health–the missing dimension in quality of life research. *Journal of pediatric rehabilitation medicine*, *12*(4), 345-359.  (Castillo *et al.*, 2019) | USA | **Type of study:** Systematic review | 18 papers included in the systematic review | In this systematic review quality of life measures were highlighted, but the missing dimension was quality of life measures in primary language. | 12 (67%) of the 18 papers reported demographic information related to race/ethnicity. Within the 12 papers that reported this demographic information, only seven articles included Hispanics/Latinos. Of these seven, a single study reported 68.2% of its participants as being Hispanics/Latinos however, the remaining studies reported a smaller proportion of Hispanics/Latinos as participants.  As a starting place, the authors would encourage the SB research community to include Hispanics/Latinos in quality of life studies and conduct interviews and questionnaires in their primary language as an estimated 55 million Hispanics/Latinos live in the USA. | A high proportion of the included papers included information on ethnicity.  The authors noted that quality of life measures were not in the users primary language in many cases, and that this should be addressed in future research.  The authors noted that future quality of life research should not only take preferred language into account, but it should also explore other dimensions keenly relevant to immigrants such as levels of family support, acculturation, adverse childhood experiences, and resilience. |
| Chowdhury, N., Naeem, I., Ferdous, M., Chowdhury, M., Goopy, S., Rumana, N., & Turin, T. C. (2021). Unmet healthcare needs among migrant populations in Canada: Exploring the research landscape through a systematic integrative review. *Journal of immigrant and minority health*, *23*(2), 353-372.  (Chowdhury *et al.*, 2021) | Canada | **Type of study:** Systematic review  **Description:** Original articles that studied unmet healthcare needs among immigrants, refugees, and/or temporary migrants in Canada were included. | 31 studies were included in this systematic review. | The systematic review was conducted to (1) identify the literature on unmet healthcare needs among different migrant populations in Canada, and (2) compile the reported factors associated with these unmet needs in various migrant groups. | Thirty-one studies reported unmet healthcare needs among migrants in Canada. The authors found five categories of unmet needs across different groups of migrants including immigrants, refugees, and temporary migrants. Immigrants and refugees face unique factors that influence the development of unmet needs, such as socio-cultural differences, communication difficulties, and lack of information. Alternatively, temporary migrants have unmet needs due to factors associated with their immigration clauses, such as healthcare coverage being conditional to work permit renewal or precarious living conditions associated with work-related housing | This was a systematic review of papers relevant to the context of Canada.  Overall, Chinese  immigrants were the most frequently studied ethnic group  (8 of 31) among all variations of samples in the different  Articles included in this systematic review. Other ethnicities studied were diverse in nature. |
| Di Pietro, N. C., & Illes, J. (2014). Disparities in Canadian indigenous health research on neurodevelopmental disorders. *Journal of Developmental & Behavioral Pediatrics*, *35*(1), 74-81.  (Di Pietro and Illes, 2014) | Canada | **Type of study:** Systematic Review  **Description:** A systematic review to map the landscape of research on autism (ASD), cerebral palsy (CP), and fetal alcohol spectrum disorder (FASD) in Canadian Aboriginal children | 52 reports published since 1981 were included in this systematic review | No specific outcome measures were utilised as this was a systematic review. | The focus on foetal alcohol spectrum disorder (FASD) in Aboriginal children and the absence of research on the other 2 major childhood disorders (CP and autism) are at odds with rates of these disorders across Canadian children. The authors argue that this trend violates fundamental principles ensuring equitable representation of all children regardless of background in research and access to benefits of research in health care and perpetuates stigma in an already marginalized population. | This systematic review highlights the paucity of research conducted with Aboriginal groups in Canada. The inclusion of these ethnic minority groups should be improved to ensure that the “views of Indian, Inuit and Métis peoples are represented in research planning and decision making, from the earliest stages of conception and design of projects through to the analysis and dissemination of results”. |
| Huang, Y., Martinez-Alvarez, M., Shallcross, D., Pi, L., Tian, F., Pan, J., & Ronsmans, C. (2019). Barriers to accessing maternal healthcare among ethnic minority women in Western China: a qualitative evidence synthesis. *Health policy and planning*, *34*(5), 384-400.  (Huang *et al.*, 2019) | China | **Type of study:** Systematic Review  **Description:** This systematic review found 10 qualitative papers covering a period during which China made substantial progress in maternal health. | 10 qualitative papers were included in this systematic review | This was a systematic review. | Poor quality of care in health facilities, particularly misunderstandings between doctors and patients due to **language barriers** or differences in socio-economic status, and clinical practices that conflicted with local fears and traditional customs, were reported. | The authors noted that eight out of 10 studies did not report the language used during interview or how responses were recorded, suggesting that this should be recorded in future studies. |
| Joo, J. Y., & Liu, M. F. (2020). Nurses’ barriers to care of ethnic minorities: A qualitative systematic review. *Western journal of nursing research*, *42*(9), 760-771.  (Joo and Liu, 2020) | Korea | **Type of study:** Systematic review  **Description:** A qualitative systematic review to identify barriers to providing healthcare services to ethnic minority patients from the perspective of nurses. | 8 papers were included in this systematic review. | This was a qualitative systematic review and no outcome measures were used. | Five common themes were identified: communication issues; unclear, missing, or culturally inappropriate care information and resources; insufficient cultural training and education; challenging therapeutic relationships with patients; and concern about quality of care. | Nurses said that it was hard to assess ethnic minority patient’ conditions and the intensity of their pain. This difficulty providing comprehensive assessment negatively influenced nurses’ ability to provide appropriate care services.  The systematic review also highlighted that on-site interpreters were not always available, and telephonic interpreters were unable to translate complex health care issues or to see facial expressions. |
| Woodall, A., Morgan, C., Sloan, C., & Howard, L. (2010). Barriers to participation in mental health research: are there specific gender, ethnicity and age related barriers?. *BMC psychiatry*, *10*(1), 1-10.  (Woodall *et al.*, 2010) | England UK | **Type of study:**  Systematic review  **Description:** Review the current literature on the nature of barriers to participation in mental health research, with particular reference to gender, age and ethnicity; b) review the evidence on the effectiveness of strategies used to overcome these barriers. | 49 papers were included in this systematic review. | No outcome measures were highlighted. | There was evidence of a wide range of barriers including transportation difficulties, distrust and suspicion of researchers, and the stigma attached to mental illness. Strategies to overcome these barriers included the use of bilingual staff, assistance with travel, avoiding the use of stigmatising language in marketing material and a focus on education about the disorder under investigation. | One of the papers in the review highlighted the need for bilingual staff as a facilitator to the barrier of language difficulties when recruiting into mental health research.  Fear, suspicion and/or distrust of researchers, concerns about confidentiality, , transportation difficulties, severity of illness, lack of financial reward, an increase in age - associated illness inconvenience and the stigma of mental illness were other barriers to taking part in mental health research. |

### Table A3 Meta analyses (n=2)

| **Citation** | **Country** | **Study details** | **Participants** | **Outcomes** | **Results** | **Overview** |
| --- | --- | --- | --- | --- | --- | --- |
| Angus, J. E., Lombardo, A. P., Lowndes, R. H., Cechetto, N., Ahmad, F., & Bierman, A. S. (2013). Beyond barriers in studying disparities in women’s access to health services in Ontario, Canada: A qualitative metasynthesis. *Qualitative health research*, *23*(4), 476-494.  (Angus *et al.*, 2013) | Canada | **Type of study:** Meta-analysis  **Description:** A meta-study approach was taken to analyse the results from 35 relevant qualitative articles to understand the conditions and conceptualizations of women’s inequitable access to health care. | 35 qualitative studies were included in this meta analysis. | A meta-analysis looking for barriers to studying health disparities. No outcome measures used. | 11 of the 35 papers referred to linguistic or cultural constraints.  Some authors cited examples of Vietnamese and Chinese immigrant new mothers who did not fully benefit from English language programs and services that were not aligned with their cultural backgrounds. | This was a meta-analysis of 35 qualitative papers investigating access to services by women in Canada.  Women valued providers who were culturally sensitive to and astute concerning issues of heteronormativity, ableism, or racism, and they appreciated the opportunity to use trained and confidential interpreters or teletype communication (TTY) in encounters with health care providers. |
| Clarke, A. R., Goddu, A. P., Nocon, R. S., Stock, N. W., Chyr, L. C., Akuoko, J. A., & Chin, M. H. (2013). Thirty years of disparities intervention research: what are we doing to close racial and ethnic gaps in health care?. *Medical care*, *51*(11).  (Clarke *et al.*, 2013) | USA | **Type of study:** Meta analysis  **Description:** A review of 11 systematic reviews. | 11 systematic reviews were included in this meta analysis.  N=391 articles in total (including 2,420 instances of tactic-level combinations) | Clarke et al (2013) used qualitative theme analysis to develop a taxonomy of disparities interventions and categorized the 391 articles accordingly. Qualitative theme analysis is a widely-used methodology for developing taxonomies in health services research. The taxonomy consisted of three components: the tactic, or what was done to intervene (e.g., communication-skills training), the strategy, or a group of tactics sharing common characteristics (e.g., delivering education and training), and the level, or who or what was targeted by the effort (e.g., the provider) | The most common strategy to improve minority health was delivering education and training (37%). The most common tactic was delivering education about a disease (14%), followed by education in disease self-management (11%). Training in communication-skills (3%) and use of decision-making aids (1%) were less frequent education tactics. The least common strategies were providing financial incentives and enhancing language and literacy services. Only one study in our review used pay-for performance to improve minority health (< 0.1%) and four offered incentives to reward healthy behaviour (0.2%). Tactics related to language such as health literacy screening and enhanced interpreter services accounted for 0.1% and 0.3% of tactics, respectively. | The least common strategy in the review of tactics/incentives were providing financial incentives and enhancing language and literacy services.  These are defined as:  Enhanced interpreter services: improvements in the quality of existing language interpreter services or the introduction of new language interpreter services.  Health literacy screening: form or interview used to gauge an individual's ability to read, understand and use healthcare information to make decisions and follow instructions for treatment. |

### Table A4 Qualitative studies (n=22)

| **Citation** | **Country** | **Study details** | **Participants** | **Outcomes** | **Results** | **Overview** |
| --- | --- | --- | --- | --- | --- | --- |
| Akhavan, S., & Karlsen, S. (2013). Practitioner and client explanations for disparities in health care use between migrant and non-migrant groups in Sweden: a qualitative study. *Journal of Immigrant and Minority Health*, *15*(1), 188-197.  (Akhavan and Karlsen, 2013) | Sweden | **Type of study:** A qualitative study  **Description:** evidence collected during in-depth interviews with health service clients and physicians. | 5‘migrant’ health service clients and 5 physicians. | Interview questions to investigate health disparities between migrant and on-migrant groups. | Explanations for disparities in health care use in Sweden can be categorized into those reflecting social/structural conditions and the presence/ absence of power and those using cultural/behavioural explanations. The negative perceptions of ‘migrant’ clients held by some Swedish physicians place the onus for addressing their poor health with the clients themselves and risks perpetuating their health disadvantage. The power disparity between doctors and ‘migrant’ patients encourages a sense of powerlessness and mistreatment among patients. | This was a qualitative study. In the findings it was noted: Language issues associated with the diagnostic instruments used in Swedish health care were also described as a factor that may lead to inequalities in health and health care. For example, physicians were aware that the questionnaires used to diagnose patients with psychological illnesses were culturally bound: ‘‘We need better tools with easier questions… You might not get this score [associated with certain diagnostic categories] from someone who does not know the nuances (nyanser)’’. |
| Betancourt, T. S., Frounfelker, R., Mishra, T., Hussein, A., & Falzarano, R. (2015). Addressing health disparities in the mental health of refugee children and adolescents through community-based participatory research: A study in 2 communities. *American journal of public health*, *105*(S3), S475-S482.  (Betancourt *et al.*, 2015) | USA | **Type of study:** A qualitative study  **Description:** research methods were used to develop community needs assessments and identify local terms for child mental health problems among Somali Bantu and Bhutanese refugees in Greater Boston and Springfield, Massachusetts, between 2011 and 2014. | **Free List Group**  N=39 = Somali Bantu  N=62 = Bhutanese refugees  **Key Informant** Group  N=21 = Somali Bantu  N=40 = Bhutanese refugees | The authors sought a broad understanding of the problems, strengths and resources of the refugee community. | The most frequently cited problems were related to language barriers (83%), including parents and children being unable to communicate with teachers and other school personnel in the USA.  When asked what helped children with these kinds of problems, the Somali Bantu refugee community was frequently identified, with 33% of participants reporting **community support as a major protective factor.** Both youths and adults reported that school personnel and parents worked together to help children with their problems. | This qualitative study addressed the health disparities of refugee children and young people in the USA. The most frequently cited problems were related to language barriers (83%), which were overcome with the help of school personnel and parents |
| Claydon‐Platt, K., Manias, E., & Dunning, T. (2014). The barriers and facilitators people with diabetes from a non-English speaking background experience when managing their medications: a qualitative study. *Journal of clinical nursing*, *23*(15-16), 2234-2246.  (Claydon-Platt, Manias and Dunning, 2013) | Australia | **Type of study:** A qualitative study  **Description:** Participants from non-English speaking backgrounds were interviewed using a semi-structured interview guide. All interviews were audio-recorded, transcribed verbatim and analysed using a thematic framework method. | 11 people with diabetes, 10 carers and 10 health professionals were interviewed  N= 31 in total | The aim of this study was to explore the barriers to and facilitators of effective medication management by people with diabetes from people from non-English speaking backgrounds from the perspectives of people with diabetes from non-English speaking backgrounds carers and health professionals. | A number of barriers were identified: cost of medications, language barriers that hinder communication, forgetfulness, and poor knowledge and understanding of medications and diabetes; however, only a small number of people used decision support tools (facilitators) such as a medication list to manage their medications | Language barriers undermined effective medication management by reducing the quantity and quality of information conveyed to people with diabetes. Likewise, poor communication resulted in non-adherence and, consequently, medication related problems.  People with low health literacy often rely on family members or friends to read and relay medication directions (Stewart et al. 1999).  A number of people with diabetes relied on their carers to relay or translate information.  It was a fact that the participants had to be able to speak English without the aid of an interpreter to take part in the study, and they still preferred to speak their native tongue, despite living in Australia for a mean time of over 44 years. |
| De La Torre, N. (2010). Hispanics with severe mental health disorders: A phenomenological study of the concern and needs of hispanics psychiatric outpatient treatment (Order No. AAI3391462). Available from APA PsycInfo®. (815572396; 2010-99180-040). Retrieved from <https://search-proquest-com.ezproxy.bangor.ac.uk/dissertations-theses/hispanics-with-severe-mental-health-disorders/docview/815572396/se-2?accountid=14874>  (De La Torre, 2009) | USA | **Type of study:** A qualitative study (PhD thesis)  **Description:** A phenomenological study of the concern and needs of Hispanic patients having psychiatric outpatient treatment | Participants included 20 Hispanic adults of both sexes (10 males and 10 females) that: (a): have been residing in the Miami-Dade County for at least 2 years, (b) have an Axis 1 diagnosis of severe mental health disorders, (c) are in the initial process of remission without periods of deterioration in the last 6 months, and (d) have been receiving psychiatric outpatient treatment for at least 1 year. These Hispanic adults with severe mental health disorders ranged in age from 21 to 55 years and were from Hispanic populations such as Mexicans (4), Cubans (4), Puerto Ricans (4), Central Americans (4), and South Americans (4). Such populations or Hispanic subgroups were selected because they are the subgroups that have more individuals living in the Miami-Dade County, as the literature reflects. | No outcome measures, but looking at medication compliance, psychiatric treatment and recovery from severe mental health disorder. | Language barriers constituted an obstacle to treatment progress because it triggered poor relationships with the psychiatrist, poor quality of care, participants’ inability to appropriately express their feelings and needs, poor communication between participants and their psychiatrist, and at times, participants’ early termination of treatment | This was a qualitative study including powerful quotes such as:  She does not speak Spanish. I would like someone that I could share my problems and feelings in my language. It is hard for the doctor to understand what I am saying due to my accent…. I would like to have a real conversation. I would like to speak with someone that is really interested in my problems, fears, and anxieties.  Also, one female respondent complained about the **lack of rapport** that she had with her psychiatrists. She did not speak Spanish; thus, the respondent needed a translator in each appointment. The respondent claimed that the psychiatrist **“was not sensitive”** to her limited English skills, and sometimes made her **“feel uncomfortable.”** |
| Dingoyan, D., Schulz, H., & Mösko, M. (2012). The willingness to participate in health research studies of individuals with Turkish migration backgrounds: barriers and resources. *European Psychiatry*, *27*(S2), S4-S9.  (Dingoyan, Schulz and Mosko, 2012) | Germany | **Type of study:** A qualitative study (focus group methodology).  **Description:** Four semi- structured focus groups of individuals with Turkish migration backgrounds living in Germany were conducted to identify potential participation barriers. | The number of participants varied between 7 and 12 individuals per focus group. | No outcome measures were used in the focus group interviews. | The authors noted that “language barriers are of relevance with regard to recruitment processes and interviews”. (P. S9).  The findings on the perception of the focus groups about how health research is perceived by individuals with Turkish migration backgrounds and the implications for successful recruitment offer considerable recommendations for enhancing participation rates in further research. Feelings of mistrust and anxiety were embedded in negative experiences with German public authorities and interactions with Germans in daily life. Against this background, it is difficult but necessary for the research staff to build trust. In addition to a field team that should include members of the target population. | The focus groups with Turkish women were not conducted in the Turkish language, but the authors noted that....  “It was also repeatedly stated that the interviewer should be able to speak the Turkish language” (P. S8). |
| Doyle, E., Rager, R., Bates, D., & Cooper, C. (2006). Using community-based participatory research to assess health needs among migrant and seasonal farmworkers. *American Journal of Health Education*, *37*(5), 279-288.  (Doyle *et al.*, 2013) | USA | **Type of study:** A qualitative study  **Description:** 5 qualitative interviews with migrant and seasonal farm workers and social and healthcare providers | N=9 Healthcare providers  N=11 Social service providers  N=20 migrant and seasonal farm workers  N = 40 in total | No specific outcome measures were utilized as this was a qualitative study. | A preliminary comparison of responses across stakeholder groups, known as heterogeneity sampling, revealed differing perspectives related to contributing factors and potential solutions. A discussion of the community-based participatory research (CBPR) process and results, and specific intervention recommendations are provided. | Although not the main focus of the investigation, the social service providers who were interviewed, pointed out service gaps in prenatal, vision, and hearing care, and a lack of Spanish-language health care information for the migrant workers in Texas, USA. |
| Fisher, C. (2011). Implications of participation and equality in the research process for health promotion practice: domestic violence as an example. *Health Promotion Journal of Australia*, *22*(2), 119-123.  (Fisher, 2011) | Australia | **Type of study:** A qualitative study  **Description:** In-depth interviews and focus groups | N=54 from five communities in Australia.  N = 24 from health and support agency staff who provide services to them.  Agency support staff represented a range of professional perspectives.  N=78 in total | No outcome measures were used, but Interviews were conducted in English, Dinka, Madi, Arabic, Somali, Krio and Amharic.  Focus groups were undertaken in English. | Community strength and capacity can be supported through research and is relevant for health promotion practice.  Being able to opt to be interviewed and give verbal consent to participate in a language other than English, facilitated participation by lay community members. The conversational nature of the interviews also facilitated lay participation and acted as an empowering agent for participants through being seen as having expertise in the issue under discussion. | This qualitative study utilized different community languages in the interviews, but not in the focus groups, the reason for this is not stated. |
| French, C., & Stavropoulou, C. (2016). Specialist nurses’ perceptions of inviting patients to participate in clinical research studies: a qualitative descriptive study of barriers and facilitators. *BMC medical research methodology*, *16*(1), 1-12.  (French and Stavropoulou, 2016) | UK | **Type of study:** A qualitative study  **Description:**  A qualitative descriptive study was conducted between March and July 2015.  Semi-structured interviews to gain a descriptive overview of barriers and facilitators. | Participants were 12 specialist nurses representing 7 different clinical specialties and 7 different NHS Trusts. | No outcome measures were utilised in this qualitative study. | Barriers and facilitators were complex and interdependent. Perceptions varied among individuals, however barriers and facilitators centred on five main themes: i) assessing patient suitability, ii) teamwork, iii) valuing research, iv) the invitation process and v) understanding the study. Facilitators to inviting patients to participate in research often stemmed from specialist nurses’ attitudes, skills and experience. Positive research cultures, effective teamwork and strong relationships between research and clinical teams at the local clinical team level were similarly important. Barriers were reported when specialist nurses felt they were providing patients with insufficient information during the invitation process, and when specialist nurses felt they did not understand studies to their satisfaction. | Strong local research culture needed to recruit participants into studies.  This study highlights important facilitators stemming from the attitudes, experience, skills and knowledge that specialist nurses bring to the invitation to clinical trials process. |
| Gaston-Johansson, F., Hill-Briggs, F., Oguntomilade, L., Bradley, V., & Mason, P. (2008). Patient perspectives on disparities in healthcare from African-American, Asian, Hispanic, and Native American samples including a secondary analysis of the Institute of Medicine focus group data. *Journal of National Black Nurses' Association: JNBNA*, *18*(2), 43-52.  (Gaston-Johansson *et al.*, 2008) | USA | **Type of study:**  A qualitative study  **Description:**  6 focus groups (secondary data) | 9 participants in each of the 6 focus groups  Ethnicity groups included African American  Chinese  Taiwanese  El Salvadoran  Cuban  Mexican  Sab Felipe (Pueblo tribe)  N=42 | No outcome measures were used in this qualitative study. | Six themes were identified that highlight patients concerns:   1. Process and resources for informed decision making in selecting providers. 2. Provider training and characteristics for cultural competence. 3. Service delivery and medical setting efficiency. 4. Treatment setting and physical enviroment. 5. Alternate models of service delivery. 6. Evaluation and oversight   The barriers and priorities for action included difficulty in making informed choices when identifying and selecting providers, poor service delivery from medical office staff, the inefficiency of medical visits, **provider communication and cultural competence barriers,** and stressful treatment settings. | In this paper, there were 6 main patient recommendations to improve minority healthcare quality, these included cultural competence including language.  The authors suggest that more ethnic minority (Spanish speaking in this instance) speakers and bilingual providers should be trained to provide a good health service for minority language speakers. |
| Haley, S. J., Southwick, L. E., Parikh, N. S., Rivera, J., Farrar-Edwards, D., & Boden-Albala, B. (2017). Barriers and strategies for recruitment of racial and ethnic minorities: perspectives from neurological clinical research coordinators. *Journal of racial and ethnic health disparities*, *4*(6), 1225-1236.  (Haley *et al.*, 2017) | USA | **Type of study:** A qualitative study  **Description:** Two semi-structured focus groups were conducted with a purposive sample of 29 clinical research coordinators (CRCs) at consecutive international stroke conferences in 2013 and 2014 to gain in-depth understanding of coordinator-level barriers to racial-ethnic minority recruitment and retention into neurological trials. | 29 clinical research coordinators (CRCs) | This was a qualitative study and no outcome measures were used. | Barriers related to translation, literacy, family composition and severity of medical diagnosis were identified. Potential strategies included a focus on developing personal relationships with patients, community and patient education, centralized clinical trial administrative systems, and competency focused training and education for clinical research coordinators (CRCs). | This was a qualitative study investigating barriers and strategies for recruitment of racial and ethnic minorities: perspectives from neurological clinical research coordinators in the USA.  Clinical Research Coordinatorss identified numerous recommendations to improve study recruitment including the need for: advanced preparation, patient education, community education, relationship building with patients and hospital staff, as well as improved CRC hiring using recommended competency assessments, and training.  Low literacy levels was perceived as the main barrier to recruitment (not just translation concerns). |
| Hunter-Adams, J., & Rother, H. A. (2017). A qualitative study of language barriers between South African health care providers and cross-border migrants. *BMC health services research*, *17*(1), 1-9.  (Hunter-Adams and Rother, 2017) | South Africa | **Type of study:**  A qualitative study  **Description:** Semi-structured in-depth interviews and focus groups with people living in Cape Town, South Africa. | Semi-structured interviews:  Congolese (n = 7)  Somali (n = 8) Zimbabwean (n = 8) women living in Cape Town  9 Focus groups including men and women. | This was a qualitative study and no outcome measures were used. | There were challenges in communication because of a lack of common language.  For example it was reported that participants’ perception of language as a vehicle of discrimination was exacerbated by participants’ experiences of discrimination outside the healthcare system. For example, among Zimbabweans, participants’ interpreted the language barrier as an unnecessary imposition: Nurses could speak English but tended to communicate in IsiXhosa, despite the fact that Zimbabweans could not understand this South African language: | In low- and middle-income (LMIC) settings health communication tends to have low priority relative to other pressing issues in the health system.  There was no effort to link patients with health care providers which shared the same language (or second language) as them, and because of this many patients felt discriminated against. |
| Johnsen, H., Kivi, N. G., Morrison, C. H., Juhl, M., Christensen, U., & Villadsen, S. F. (2020). Addressing ethnic disparity in antenatal care: a qualitative evaluation of midwives’ experiences with the MAMAACT intervention. *BMC pregnancy and childbirth*, *20*(1), 1-10.  (Johnsen *et al.*, 2020) | Denmark | **Type of study:** A qualitative study (mini group interviews).  **Description:** Eight mini-group interviews with midwives (n = 18) were undertaken to explore the feasibility of the MAMAACT intervention, which included a training course for midwives, a leaflet and a mobile application, as well as additional visit time, was developed and tested at a maternity ward to increase responses to pregnancy warning signs among midwives and non-Western immigrant women. | N=18 Midwives | This was a qualitative study and no outcome measures were used. | Three main categories were identified, which were ‘Challenges of working with non-Western immigrant women’, ‘Attitudes towards and use of the leaflet and mobile application’, and ‘Organisational factors affecting the use of the MAMAACT intervention’. | The authors noted that language proficiency was of great importance for the provision of care. Midwives had concerns about communication difficulties causing adverse events. Many non-Western immigrant women were described as lacking the ability to express themselves in Danish or English. Even though the hospital offered interpreter assistance, interpreters were not always available for the midwifery visits. Sometimes, immigrant women would bring their partner, a relative or a friend to interpret for them. This was described as potentially problematic due to the lack of confidentiality and the ability to assess the quality of the translation. Midwives could be uncertain if women’s symptoms were described accurately and if their information and advice were conveyed as intended. In situations where no interpreters or family members were available to interpret, midwives would try to get by using gestures or simple words to assess the health of the mother and the baby. |
| MacFarlane, A., Singleton, C., & Green, E. (2009). Language barriers in health and social care consultations in the community: a comparative study of responses in Ireland and England. *Health Policy*, *92*(2-3), 203-210.  (MacFarlane, Singleton and Green, 2009) | Ireland and England | **Type of study:** Comparative analysis  **Description:** Comparative analysis of two action research studies (one in Ireland and one in England). | **Ireland:** N=26 Serb Croat and Russian speaking refugees and asylum seekers (n = 16 females and n = 10 males).  **England:**  Focus groups (11 focus groups; n = 61 participants)  Semi-structured interviews (n = 28 participants). | This was a comparative analysis and no outcome measures were used. | Key findings are that the same range of formal and informal responses to language barriers occurs in practice in both England and Ireland, but proportions of knowledge and use of these responses differ. English service providers have more awareness about the use of formal responses than Irish service providers but uptake of formal responses remains low in both England and Ireland.  Data from service users confirms these findings.  There is a need for more attention to the implementation of policies for language barriers in both Ireland and England, further research about the normalization processes associated with these consultations and knowledge transfer networks to facilitate on-going dialogue between all key stakeholders with an emphasis on supporting service users’ involvement and participation | There is a need for more attention to the implementation of policies for language barriers in both Ireland and England, further research about the normalization processes associated with these consultations and knowledge transfer networks to facilitate on-going dialogue between all key stakeholders with an emphasis on supporting service users’ involvement and participation.  Example comments from service users:  CARe Z6 explained that his wife likes him to interpret to give her confidence and support when describing her problems.  CARe Z3, who takes her daughter to her GP consultations to interpret said that she does not go to see the doctor if the complaint is of a personal nature that she does not want her daughter to be involved in. |
| O'Connor, M. R., Adem, A., & Starks, H. (2018). East African perceptions of barriers/facilitators for pediatric clinical research participation and development of the inclusive research model. *Journal of pediatric nursing*, *42*, 104-110.  (O’Connor, Adem and Starks, 2018) | USA | **Type of study:** A qualitative study  **Description:** Community leader interviews (n = 6) and focus groups with lay members (n = 16) from the three largest East African communities in the Seattle area (Eritrean, Ethiopian and Somali) were conducted. Discussions were semi-structured based on existing barrier/facilitator research and analyzed using directed content analysis to identify major themes | Community leader interviews (n = 6) and focus groups with lay members (n = 16) from the three largest East African communities in the Seattle area (Eritrean, Ethiopian and Somali) were conducted. | No outcome measures were used in the interviews or focus group interviews. | Community leader interviews (n = 6) and focus groups with lay members (n = 16) from the three largest East African communities in the Seattle area (Eritrean, Ethiopian and Somali) were conducted. | To facilitate more inclusive research participation, researchers, nurses and other health care providers might consider ensuring adequate time for discussion of the research study and process, engaging the community in the research process, employing lay reviews of translated materials and/or oral consent processes, and other strategies outlined in the Inclusive Research Model. |
| Robinson, J. M., & Trochim, W. M. (2007). An examination of community members’, researchers’ and health professionals’ perceptions of barriers to minority participation in medical research: an application of concept mapping. *Ethnicity and Health*, *12*(5), 521-539.  (Robinson and Trochim, 2007) | USA | **Type of study:** A qualitative study  **Description:** Focus group methodology.  A structured form of concept mapping (Trochim 1989) was the methodology used in this study. The concept mapping process has three specific phases: (1) project planning*development of project focus statements and sample selection (2) idea generation and structuring and (3) analysis and interpretation. | Across these 14 networks, a convenience sample consisting of steering committee members (n =20), community advisory board members (n =16), regional advisory board members (n=6), and lay community members (n=5) was utilized | No outcome measures were used in this qualitative study | There appears to be a difference in the barriers to participation as defined by community members themselves, and health professionals’ perceptions of these barriers. | This paper highlighted the need to record why some potential participants decline to take part in research.  Increased inclusion of minorities in the design, management, and implementation of medical research studies would help mitigate negative perceptions of the research process, and serve to increase participation among racial/ethnic minorities. |
| Sadavoy, J., Meier, R., & Ong, A. Y. M. (2004). Barriers to access to mental health services for ethnic seniors: The Toronto study. *The Canadian Journal of Psychiatry*, *49*(3), 192-199.  (Sadavoy, Meier and Ong, 2004) | Canada | **Type of study:**  A qualitative study (Action Research)  **Description** An action-research project used qualitative methodology based on grounded theory to generate areas of inquiry. | N = 17 focus groups | No outcome measures were used in this qualitative study. | The dearth of appropriate psychiatrists with language and cultural competency is the most clearly identified gap in mental health service provision. | There is a lack of psychiatrists to provide therapeutic mental health services for ethnic seniors in Toronto, Canada.  Culturally derived beliefs complicate the reactions of seniors and families to emotional distress and probably lead to delays in seeking help for mental health issues.  Key service provided attributes included being:  Trustworthy Respectful Empathetic Taking enough time Willing to talk (for example, engage with client) Understanding Awareness of mental health issues. |
| Schildmann, E. K., Groeneveld, E. I., Denzel, J., Brown, A., Bernhardt, F., Bailey, K., ... & Murtagh, F. E. (2016). Discovering the hidden benefits of cognitive interviewing in two languages: The first phase of a validation study of the Integrated Palliative care Outcome Scale. *Palliative medicine*, *30*(6), 599-610.  (Schildmann *et al.*, 2016) | Germany and England | **Type of study:**  A qualitative study  **Description:** Bi-national (United Kingdom/Germany) cognitive interview study using ‘think aloud’ and verbal probing techniques. | A total of 15 German and 10 UK interviews were conducted. | The Integrated Palliative care Outcome Scale was refined by consensus following the cognitive interviews. | Cognitive interviewing proved valuable to increase face and content validity of The Integrated Palliative care Outcome Scale. The concurrent approach in two languages benefited the refinement. | Face and content validity of an outcome measure can be enhanced through testing more than one language at the same time. In this instance German and English (only). |
| Shattell, M. M., Hamilton, D., Starr, S. S., Jenkins, C. J., & Hinderliter, N. A. (2008). Mental health service needs of a Latino population: A community-based participatory research project. *Issues in mental health nursing*, *29*(4), 351-370.  (Shattell *et al.*, 2008) | USA | **Type of study:**  A qualitative study  **Description:** A community-based participatory research project  Data were collected from October 2006 through February 2007. The CBPR team functioned as a focus group, and four two-hour focus groups were held over four months. | N = 7 community members (2 x male and 5 x female)  N = 1 health educator  N = 1 doctoral student in Nursing  N = 2 undergraduate nursing students  N = 1 Principal investigator | No outcome measures were used in this qualitative study. | Mental health service needs were discussed by members of the community research team and appropriate solutions were proposed. These mental health service needs and proposed solutions are presented below at the individual, organization, and community levels of the Social Ecological Model. | The socio-ecological model was described on P.354, and refers to solutions at the individual, organisation and community levels.  Individual level example included language: Inability to speak English was seen as a barrier to job attainment and ability to access health education messages and mental health services P.358.  Even when an American provider spoke Spanish, the Latinos were apprehensive about sharing their health concerns and viewed them suspiciously; “I meet with somebody; they’re always kind of guarded because they’re wondering ‘What is this American doing in my house? And how is it they speak Spanish? And what’s going on?”’ (P.358).  Organisational level example: There are only a few bilingual (healthcare) providers, therefore it is hard to refer.  “If you get a Spanish-speaking person in, you are really limited as to who you can refer them to.” P. 361  Community level example: One of the greatest assets for the Latino community is its own people. As this CBPR group demonstrated, community members understand their own problems. Moreover, communities provide support and information to newcomers on issues ranging from where to live, bank, and shop to how to access medical and mental health services. P.362. (e.g. churches, schools public libraries, and community centres). |
| Squires, A., Miner, S., Liang, E., Lor, M., Ma, C., & Stimpfel, A. W. (2019). How language barriers influence provider workload for home health care professionals: A secondary analysis of interview data. *International journal of nursing studies*, *99*, 103394.  (Squires *et al.*, 2019) | USA | **Type of study:** A qualitative study  **Description:** A qualitative secondary data analysis using a summative content analysis approach was used to analyse existing semi-structured interview data. | 35 home health care providers [31 registered nurses, 3 physical therapists, 1  occupational therapist]. | No outcome measures were used in this qualitative study. | The results included details regarding:  Barriers included the effort of getting in touch with interpreters  Irritation of having to wait for interpreters (patients and nurses)  Paying for interpreters (sometimes the service is there depending on demand) sometimes the health insurance covers it, but not in every state).  Problems with interpreter service including threat to patient safety due to accuracy of interpretation in healthcare settings.  In the United States, access to interpreter services is a civil right and individuals may take legal action through anti-discrimination lawsuits if language access services were implemented insufficiently to meet their communication needs. With demand for interpreter services increasing annually due to demographic changes where now one in five household in the United States speaks a language other than English at home, risks to healthcare organizations for lawsuits will increase unless language access services are implemented more systematically. | This study provides a foundation for examining the intersections of how language barriers may affect quality of care in home health care and the relationship to provider workloads. |
| Strohschein, F. J., Merry, L., Thomas, J., & Gagnon, A. J. (2010). Strengthening data quality in studies of migrants not fluent in host languages: a Canadian example with reproductive health questionnaires. *Research in nursing & health*, *33*(4), 369-379.  (Strohschein *et al.*, 2010) | Canada | **Type of study:**  A qualitative study  **Description:** Linguistic validation of different language versions of a questionnaire about reproductive health (used in Canada).  Interviews concerning the meaning, clarity, and relevance of instruments. | For the PACBIRTH study, five refugee/asylum seeking monolingual women per language, speaking Hindi, Tamil, Urdu, Spanish, and French participated in the testing. For the KAP study, three monolingual French-speaking and refugee/ asylum-seeking women participated, as well as 10 refugee/asylum-seeking monolingual Urdu-, Tamil-, and Hindi-speaking couples. Monolingual participants came from Mexico, Peru, Colombia, Pakistan, Sri Lanka, India, Cameroon, and the Congo, and were Muslim, Christian, and Hindu. | A reproductive health questionnaire was linguistically validated into several different languages. | Questionnaire revisions were made on the basis of the feedback form the interviews/group interviews.  Reproductive health questionnaires were tested with persons monolingual in Hindi, Tamil, Urdu, Spanish, and French. | The authors highlight the need to be culturally as well as linguistically aware.  Familiarity of the research process is influenced by cultural background. |
| Tatari, C. R., Andersen, B., Brogaard, T., Badre-Esfahani, S. K., Jaafar, N., & Kirkegaard, P. (2020). Perceptions about cancer and barriers towards cancer screening among ethnic minority women in a deprived area in Denmark–a qualitative study. *BMC Public Health*, *20*(1), 1-10.  (Tatari *et al.*, 2020) | Denmark | **Type of study:**  A qualitative study  **Description:**  Individual and group interviews  Perceptions about cancer and barriers towards cancer screening among ethnic minority women in a deprived area in Denmark– | A total of 37 women from ten different non-Western countries participated in the study based on five semi-structured focus groups, two structured group interviews with an interpreter and three individual interviews with culture experts. | No outcome measures were used in this qualitative study. | Cancer was perceived as a deadly disease that could not be treated. Cancer screening was perceived as only relevant if the women had symptoms. Knowledge about cancer screening was fragmented, often due to inadequate Danish language skills and there was a general mistrust in the Danish healthcare system due to perceived low medical competences in Danish doctors. There was, however, a very positive and curious attitude regarding information about the Danish cancer screening programmes and a want for more information. | Inadequate language skills in Danish was one of the four themes emerging.  "... So all the material you get you don’t read it because you don’t understand it … If it is only in Danish, no one looks at it". (P. 6 of 10).  Some communities depend on the spoken word, rather than the written word The participants reported that the Arab and Somali communities in Denmark are verbal cultures, and some of the respondents said that they get lots of information by contacting others on social media. |
| Vandan, N., Wong, J. Y. H., Lee, J. J. J., Yip, P. S. F., & Fong, D. Y. T. (2020). Challenges of healthcare professionals in providing care to South Asian ethnic minority patients in Hong Kong: A qualitative study. *Health & social care in the community*, *28*(2), 591-601.  (Vandan *et al.*, 2020) | Hong Kong, China | **Type of study:**  A qualitative study  **Description:**  Interviews with healthcare professionals | N = 22 Health care professionals | No outcome measures were used in this qualitative study | Language barriers were identified of the analysis e.g. One doctor said “There are no health education talks in South Asian languages about disease and its management in hospitals, such as after surgery. I think that health talks in South Asian common languages such as Hindi will be useful, or using large fonts and picture posters can be beneficial to impart education to SA patients”. P. 596 | This qualitative study highlighted the insufficient use of interpretation services, despite their availability. Participants raised concerns about the quality and timeliness of interpretation services and recommended on-site interpreters at hospital pharmacies as a starting point to overcome communication barriers.  The study highlighted the challenges healthcare professionals face due to inadequate cultural competency training and education provision when considering caring for South Asian patients in Hong Kong. |

### Table A5 Cohort study (n=1)

| **Citation** | **Country** | **Study details** | **Participants** | **Outcomes** | **Results** | **Overview** |
| --- | --- | --- | --- | --- | --- | --- |
| Blanchet, R., Sanou, D., Nana, C. P., Pauzé, E., Batal, M., & Giroux, I. (2017). Strategies and challenges in recruiting black immigrant mothers for a community-based study on child nutritional health in Ottawa, Canada. *Journal of immigrant and minority health*, *19*(2), 367-372.  (Blanchet *et al.*, 2017) | Canada | **Type of study:** A quantitative cohort study.  **Description:** A quantitative exploratory study was conducted to identify barriers to participation as well as recruitment strategies to engage minority parents of young children in health-oriented research. | N=259 Parent and child dyads (251 biological mothers, one adoptive mother, three fathers). | Recruiting strategies were analysed for this paper. No outcome measures were used. | The authors found that direct contact between participants and research team members (e.g. during community events) as well as referrals by someone they trusted (e.g. a friend, community partners) were the most effective recruiting strategies.  Most recruiters were Francophones, and this probably explains they recruited more Francophone than Anglophone mothers. | The quantitative study recruitment methods showed that it is important to focus on recruiter attributes, including languages, when recruiting for a study in community events. |

### Table A6 Survey studies (n=9)

| **Citation** | **Country** | **Study details** | **Participants** | **Outcomes** | **Results** | **Overview** |
| --- | --- | --- | --- | --- | --- | --- |
| Brisset, C., Leanza, Y., Rosenberg, E., Vissandjée, B., Kirmayer, L. J., Muckle, G., ... & Laforce, H. (2014). Language barriers in mental health care: A survey of primary care practitioners. *Journal of immigrant and minority health*, *16*(6), 1238-1246.  (Brisset *et al.*, 2014) | Canada | **Type of study:** A quantitative study (survey)  **Description:** A self-administered survey was developed to address their experiences of working with people neither first language English or French in Canada (allophones). | 113 primary care practitioners in Montreal. | This was a self-administered questionnaire design. No outcome measures were used. | Having access to interpreters was considered as the most important resource to overcome language barriers, but the great majority of practitioners had not been trained to work with interpreters (ad hoc or professional).  Most interpreted consultations involved ad hoc interpreters drawn from the client’s family members or friends. This finding is consistent with the existing literature ad hoc interpreters offer the advantage of immediate availability (being present at the same time as the client), continuity (being present for each consultation), trust by clients and they do not necessarily convey clients’ disagreement or resistance about the diagnostic and treatment. | This was a quantitative survey of medical practitioners in Canada. Minority language speakers in Canada (not English or French as their first language) are not trained to work with interpreters.  The authors note that training should highlight the benefits and limitations of the different roles that interpreters can play in health care delivery and the differences in communication dynamics with each role. |
| Carlini, B. H., Safioti, L., Rue, T. C., & Miles, L. (2015). Using Internet to recruit immigrants with language and culture barriers for tobacco and alcohol use screening: a study among Brazilians. *Journal of immigrant and minority health*, *17*(2), 553-560.  (Carlini *et al.*, 2015) | USA | **Type of study:** A quantitative study (survey)  **Description:** This study focused on internet recruitment methods | Brazilian community members living in the USA: (Florida, California and New Jersey). | Adverts using the internet were utilised to screen for tobacco and alcohol use. No outcome measures used. | The study was advertised in Portuguese (a minority language within the USA) using Facebook, Google, online newsletters and E-mail. Participants clicked ads to consent and access a screening for tobacco and alcohol dependence. Ads yielded 690 screening responses in 90 days. Online recruitment of populations is feasible. Future studies should test similar strategies in other LEP groups. | An internet recruitment method utilizing the Portuguese language (which is a minority language in the USA) was successful in recruiting Brazilian immigrants to a study on tobacco and alcohol use. |
| Coustasse, A., Bae, S., Arvidson, C., Singh, K. P., & Trevino, F. (2009). Disparities in ADL and IADL disabilities among elders of Hispanic subgroups in the United States: Results from the National Health Interview Survey 2001-2003. *Hospital topics*, *87*(1), 15-23.  (Coustasse *et al.*, 2010) | USA | **Type of study:** Quantitative study (National survey)  **Description:** The authors applied chi-square analysis for bivariate comparisons and used multiple logistic regression analyses for making comparisons, estimating odds ratios, and predicting disabilities. | Data from the National Health Interview Survey (2001–2003; N = 31,875).  One adult from each household is selected at random and administered a health-oriented questionnaire (i.e., the adult core), which includes questions about Activities of Daily Living (ADL) and Instrumental Activity of Daily Living (IADL). | Activity of Daily Living (ADL) and Instrumental Activity of Daily Living (IADL) | Results revealed a 21.4% rate of disability of any type in Hispanics. Puerto Ricans reported the highest rates of Activity of Daily Living (ADL) and Instrumental Activity of Daily Living (IADL) disabilities compared with other Hispanic subgroups (Mexicans, Cubans, Central and South Americans) and reported a higher rate than did Blacks. Cubans showed the lowest rate of IADL and any disability among Hispanics and a lower rate than did Whites. | The authors noted that health measures in the National Health Interview Survey have not been clinically confirmed, and the **language used in the interviews may have affected respondents’ self-reported health status.**  **The authors do not offer any evidence to support this.**  For example, the authors noted that:  NHB men were 38% less likely than were NHB women to report Activities of Daily Living (ADL) disability (p < .05). |
| Falla, A. M., Veldhuijzen, I. K., Ahmad, A. A., Levi, M., & Richardus, J. H. (2017). Language support for linguistic minority chronic hepatitis B/C patients: an exploratory study of availability and clinicians’ perceptions of language barriers in six European countries. *BMC Health Services Research*, *17*(1), 1-8.  (Falla *et al.*, 2017) | Based in Netherlands, but on-line survey conducted in 6 European Countries | **Type of study:** A quantitative study (on-line survey in different languages)  **Description:** An online survey was developed, translated and sent to experts in five health care services involved in screening or treating viral hepatitis in six European countries: Germany, Hungary, Italy, the Netherlands, Spain and the United Kingdom (UK). | N=238 respondents to on-line survey. | No outcome measures in this on-line survey of language barriers in health in European countries. | Interpreters are common in the UK, the Netherlands and Spain but variable or rare in Germany, Hungary and Italy. Translated materials are rarely/never available in Hungary, Italy and Spain but commonly or variably available in the Netherlands, Germany and the UK. Differing levels of agreement that language barriers explain the three scenarios are seen across the countries. Professionals in countries with most infrequent availability (Hungary and Italy) disagree strongest that language barriers are explanations.  In the UK, over half agree or strongly agree that language barriers explain all three scenarios (screening uptake, screening offer and referral), and only a minority (between 7 and 15%) expressed disagreement (Table 2). Strongest agreement in the UK emerges about the role of language barriers in referral, where nearly three quarters (73%) strongly agree that these explain why cases do not reach secondary care. | In this on-line survey there was evidence that minority language speakers with chronic hepatitis B/C do not receive specialist care as they do not attend hepatitis screening.  For example, in Italy 80% agree that language barriers explain the lack of hepatitis screening in primary care. |
| Greene, R. A., Karavatas, S. G., Cooper, J., & Zamorano-Torres, N. (2013). Perceptions of Spanish speaking individuals regarding the impact of language barriers on physical therapy interventions: a pilot study. *Journal of the National Society of Allied Health*, *10*(1), 75-83.  (Greene *et al.*, 2013) | USA | **Type of study:** A qualitative pilot study (survey)  **Description:** The purpose of this pilot study was to determine if limited English proficiency (LEP). LEP patients were being adversely affected by the language barriers, and to identify the perceptions of Spanish speaking individuals regarding their encounters with the health care system, specifically rehabilitation services. | A convenience sample of 30 patients in the Washington, DC metropolitan area, whose primary language is Spanish, was used. The participants were recruited from three different Physical Therapy clinics in the Washington, DC metropolitan area. All of the clinics were classified as ambulatory care facilities. | The instrument of this survey was a questionnaire, which was developed by the researchers. The investigators developed a questionnaire consisting of seven demographic questions and 12 closed- end Likert questions, in order to assess the Spanish speaking patients’ perceptions about the quality of their Physical Therapy services. The surveys were written in English and Spanish. | A number of the findings of this pilot study are consistent with research literature describing the use of family members as interpreters for patients with limited English proficiency.  Although unclear from the results, it appears that at least 25% had difficulty understanding their physical therapist. The patients in this study were, in general, not negatively affected by language barriers, possibly because a number of the practitioners were Spanish speaking. Language barriers may have affected the quality of health care provided; however, most patients surveyed appeared to receive effective care. | The patients in this study were not negatively affected by language barriers, possibly because a number of the practitioners were Spanish speaking. The findings of this study cannot be generalized, because of the small sample size, and the lack of random sample selection. Additional research is required in order to assess whether the results from this study can be replicated.  The authors highlighted workforce planning – to get those skilled in Spanish and English may be more cost beneficial in Spanish speaking areas than providing translation services. |
| Hahm, H. C., Lahiff, M., Barreto, R. M., Shin, S., & Chen, W. Y. (2008). Health care disparities and language use at home among Latino, Asian American, and American Indian adolescents: Findings from the California Health Interview Survey. *Journal of Community Psychology*, *36*(1), 20-34.  (Hahm *et al.*, 2008) | USA | **Type of study:** A quantitative study (telephone Survey)  **Description:** This telephone survey study examined the association between language use at home and self-assessed health, health insurance, and having a usual source of care among California’s minority adolescents. | 2,230 ethnic minority adolescents were interviewed over the telephone. | Questions on:  health status, medical insurance, and having a usual source of care were asked as part of the 2001 California Health Interview Survey. | Adolescents who spoke exclusively another language (other than English) were more likely to report fair or poor health (OR = 2.37, p  = .012) and to not have medical insurance (OR = 4.61, p  = .001) compared to adolescents in Group 1 (English only). Young adolescents in Group 3 (other languages) were more likely to have no usual source of care (OR = 4.07, p = .029). | Delivering appropriate care to adolescents includes speaking their language. This paper recognises that language is an increasing issue in healthcare delivery due to migration.  Interviews with adolescents were conducted in six languages: English, Spanish, Chinese (Mandarin and Cantonese), Vietnamese, Korean, and Khmer (Cambodian). The choice of languages was based on research that identified these as the languages used by the most Californians who would not be able to complete a telephone interview in English.  The authors stated that Interventions that are specifically targeted to improving ethnic minorities’ access to care have been found to be effective at the individual, organizational, and community levels these include outreach activities, bilingual/bicultural staff, and collaboration with community-based organizations. |
| Kim, G., Loi, C. X. A., Chiriboga, D. A., Jang, Y., Parmelee, P., & Allen, R. S. (2011). Limited English proficiency as a barrier to mental health service use: A study of Latino and Asian immigrants with psychiatric disorders. *Journal of psychiatric research*, *45*(1), 104-110.  (Kim *et al.*, 2011) | USA | **Type of study:**  A quantitative study (survey)  **Description:** The purpose of this study was to examine the effect of limited English proficiency (LEP) on mental health service use among immigrant adults with psychiatric disorders. Drawn from the National Latino and Asian American Study (NLAAS), Latino and Asian immigrant adults. | N=372 in total (Latino, Hispanic and Asian adults). | English-speaking ability was assessed using a single question “How well do you speak English?” Responses were dichotomized into “excellent/good (coded as 1)” or “fair/poor (coded as 0).” Those who reported their English-speaking ability as fair/poor were deemed to have limited English proficiency (LEP).  Mental health was also self-rated. | Limited English proficiency was a barrier to mental health service use among Latino immigrants with psychiatric disorders. This study suggests that future approaches to interventions might be well advised to include not only enhancing the availability of bilingual service providers and interpretation services but also increasing awareness of such options for at least Latino immigrants | Limited English proficiency is a critical barrier to mental health services for Latino immigrants. Attention needs to be paid to ways to facilitate access to services for those populations. Emphasis should be given not only to enhancing the availability of bilingual service providers and interpretation services but also to increasing awareness of such options.  Asian people were less likely to report mental health symptoms and therefore less likely to seek access to services. More research is needed. |
| Lu, S. H., Dear, B. F., Johnston, L., Wootton, B. M., & Titov, N. (2014). An internet survey of emotional health, treatment seeking and barriers to accessing mental health treatment among Chinese-speaking international students in Australia. *Counselling Psychology Quarterly*, *27*(1), 96-108.  (Lu *et al.*, 2014) | Australia | **Type of study:** A quantitative study (on-line survey)  **Description:** An internet survey of emotional health, treatment seeking and barriers to accessing mental health treatment among Chinese-speaking international students in Australia | N = 144 respondents were Chinese-speaking international students in Australia | Kessler-10 item measure of psychological distress. | Chinese-speaking international students are a high risk group for developing psychological distress, yet they tend to underuse mental health services. Education about the effectiveness of face-to-face and online treatments may increase treatment seeking by this population. | Among those who reported high psychological distress on the K-10 and responded to the question on barriers to accessing treatment (n = 65, 45%), frequently endorsed practical barriers were the concern about treatment costs or transportation difficulties, lack of knowledge of treatment services, time constraints, difficulties and lack of knowledge of symptoms of low mood, anxiety and stress. Common cultural barriers reported by the respondents include not perceiving symptoms as severe or serious enough to warrant treatment and language difficulties. |
| Mosconi, P., Antes, G., Barbareschi, G., Burls, A., Demotes-Mainard, J., Chalmers, I., ... & Wolff, S. (2016). A European multi-language initiative to make the general population aware of independent clinical research: the European Communication on Research Awareness Need project. *Trials*, *17*(1), 1-10.  (Mosconi *et al.*, 2016) | Italy  (European countries) | **Type of study:** A quantitative study (survey)  **Description:** The authors searched for, and evaluated, relevant existing materials and developed additional materials and tools, making them freely available under a Creative Commons licence. | 1,852 survey responses. | No outcome measures were used in this survey of resources. | The principal communication materials developed were: 1. A website (http://ecranproject.eu) in six languages, including a Media centre section to help journalists to disseminate information about the ECRAN project. An animated film about clinical trials, dubbed in the 23 official languages of the European Community, and an interactive tutorial. An inventory of resources, available in 23 languages, searchable by topic, author, and media type. Two educational games for young people, developed in six languages. Testing Treatments interactive in a dozen languages, including five official European Community languages. An interactive tutorial slide presentation testing viewers’ knowledge about clinical trials | To ensure efficient dissemination of resources, all the materials developed for lay people were translated and dubbed into at least six European languages.  The inventory of ECRAM resources, is available in 23 languages, searchable by topic, author, and media type.  As the language of medical science is English, people who cannot understand English face a barrier to obtaining intelligible information. This has been partially overcome by ECRAN (P.7 of 10). |

### Table A7 Mixed method studies (n=9)

| **Citation** | **Country** | **Study details** | **Participants** | **Outcomes** | **Results** | **Overview** |
| --- | --- | --- | --- | --- | --- | --- |
| Ahlmark, N., Algren, M. H., Holmberg, T., Norredam, M. L., Nielsen, S. S., Blom, A. B., ... & Juel, K. (2015). Survey nonresponse among ethnic minorities in a national health survey–a mixed-method study of participation, barriers, and potentials. *Ethnicity & health*, *20*(6), 611-632.  (Ahlmark *et al.*, 2014) | Denmark | **Type of study:** mixed-method study.  **Description:** A logistic regression was used to analyse non-response in a national health survey using data from the Danish National Health Survey (DNHS). | Data from a large Danish National Health Survey (DNHS). (N = 177,639.  Also data from 10 immigrants and 13 descendants between the ages of 18 to 54. | Completion or non-completion of the national health survey questionnaire in Denmark. | The highest nonresponse rate was for non-Western descendants (80.0%) and immigrants 25 (72.3%) with basic education. Immigrants and descendants had higher odds ratios (OR = 3.07 and OR = 3.35, respectively) for nonresponse than ethnic Danes when adjusted for sex, age, marital status, and education. Non-Western immigrants had higher item nonresponse in several question categories. Barriers to non-participation related to the content, language, format, and layout of both the questionnaire and the cover letter. The sender and setting in which to receive the questionnaire also influenced answering incentives. The authors observed differences in barriers and incentives between immigrants and descendants. | A mixed method study of a large quantitative dataset and qualitative focus groups.  The authors noted that in present study, they did not look into the possibility of using different language versions of the questionnaire; however, this could perhaps increase the response rate among non-Danish-reading immigrants. |
| Bhuiyan, B. A., Urmi, I. J., Chowdhury, M. E., Rahman, T., Hasan, A. S., & Simkhada, P. (2019). Assessing whether medical language is a barrier to receiving healthcare services in Bangladesh: an exploratory study. *BJGP open*, *3*(2).  (Bhuiyan *et al.*, 2019) | Bangladesh | **Type of study:** A quantitative study (survey).  **Description:** A semi-structured questionnaire was developed through Google Forms for data collection. | 50 participants of different age groups.  N= 44 male  N = 6 female | A questionnaire was developed to answer the research question | Despite two languages being used in the medical context (English and Bengali) in Bangladesh, there are many minority languages, that are not used in the medical context.  There was qualitative findings to show that at least some respondents would have liked all of their medical instructions in Bengali.  Medical language is already different from the native language people speak in their day-to-day conversations.  As such, the language difference between doctors and patients sometimes becomes a strong barrier to achieving a successful treatment outcome. | This was a quantitative study assessing medical languages causing barriers to healthcare in Bangladesh.  The authors also noted that there is strong evidence from around the world that has proven that miscommunication among health providers and patients plays a major role in the healthcare system, and that medical language is acting as a barrier to achieving an effective health service. |
| Chirewa, B. (2012). Development of a practical toolkit using participatory action research to address health inequalities through NGOs in the UK: Challenges and lessons learned. *Perspectives in public health*, *132*(5), 228-234.  (Chirewa, 2012) | UK | **Type of study:** Mixed method research  **Description:** A mixed method research study within a participatory action research framework. Including semi-structured questionnaire and focus groups and discussions with experts. | The research participants were drawn from the National Government Organisations (NGO’s) forum affiliates (n = 30), NHS primary care trusts (PCTs) (n = 2) and an expert group (academics) (N = 4) with an interest in health inequalities and the third sector. | The primary outcome was to develop a practical toolkit to address health inequalities. | Recognizing, respecting and embracing the different **cultures** of the stakeholders and partner NGOs is imperative for successful participatory action research efforts. Addressing **diversity** can foster inclusiveness and active participation of members. It is therefore important to reach out to as many NGO forum members as possible, with an emphasis being placed on the very small NGO organizations in order to mirror the various subgroups working with the hard-to-reach, marginalized communities that have historically been ‘invisible’. | This was a mixed method study.  It is interesting to note that some groups declined to be involved in the study as they saw no benefit in supplying evidence when the final document would not be presented in their native language, for instance Hindi.  Despite the fact that some stakeholder groups did not take part because of language issues, the authors did not identify ‘language’ as one of their lessons learnt or as a recommendation moving forward. |
| Eriksson‐Sjöö, T., Cederberg, M., Östman, M., & Ekblad, S. (2012). Quality of life and health promotion intervention–a follow up study among newly‐arrived Arabic‐speaking refugees in Malmö, Sweden. *International Journal of Migration, Health and Social Care*.  (Eriksson-Sjöö *et al.*, 2012) | Sweden | **Type of study:** Mixed method study  **Description:** Questionnaires, observations and oral evaluations in groups | N = 78 newly‐arrived Arabic‐speaking adult refugees in Malmö, Sweden took part in one or more elements of this mixed methods study. | HRQoL was measured by EQ-5D self-assessment (EuroQoL Group, 1990), which is translated/back translated into Arabic (Lebanese version) and copyrighted. | As lack of language skills can be a major barrier to understanding bureaucratic procedures and the functioning of the health system, it is necessary that society provides sufficient numbers of adequately trained professional interpreters when needed | This mixed method study emphasised that language is an important aspect of care for newly arrived Arabic speaking refugees in Sweden.  The authors noted that as migrants face specific difficulties with respect to their right to health, it is important that education/training in medicine, health care and social work provide cultural competence. |
| Mowlabaccus, W. B., & Jodheea-Jutton, A. (2020). Participant perception, still a major challenge to clinical research in developing countries—A mixed methods study. *Plos one*, *15*(7), e0236563.  (Mowlabaccus and Jodheea-Jutton, 2020) | Mauritius | **Type of study:** Mixed method research  **Description:** A mixed study was carried out which consisted of 2 phases: a qualitative, with thematic approach followed by a quantitative study with cross-sectional design.  . | **Qualitative:** N = 23 open-ended question on-line survey.  **Survey:** N= 350 completed questionnaires | A validated questionnaire for India and Korea was adapted for use in Mauritius. | Most of the respondents agreed with the value of research while a minority had poor perception related to trust in research companies and conduct of clinical trials. Respondents who had previously engaged in clinical research had better knowledge and perception compared to those who did not participate in one. | There was a generalized vagueness with regards to the concept of clinical trials.  Informed consent should be in the language of the participant.  Literacy level should also be considered.  Regarding perception of clinical trials, and literacy level, a better perception score was found to be associated with a high literacy level (p value = 0.02). (P.6 of 15). |
| Tadić, V., Hamblion, E. L., Keeley, S., Cumberland, P., Hundt, G. L., & Rahi, J. S. (2010). ‘Silent voices’ in health services research: ethnicity and socioeconomic variation in participation in studies of quality of life in childhood visual disability. *Investigative ophthalmology & visual science*, *51*(4), 1886-1890.  (Tadić *et al.*, 2010) | UK | **Type of study:**  Mixed method study including interviews and survey  **Description:**  Mixed method including interviews and PedsQoL | N = 32 – interviewed (in stage 1)  N = 44 (in stage 2)  Different populations. | The authors examined the QoL of 44 children and adolescents with hereditary retinal disorders, which enrolled in the parent study using a generic multidimensional pediatric tool for assessing children’s health-related (HR) QoL (Pediatric Quality of Life Inventory [PedsQL 4.0]). | The overall participation level was below 50%. In both studies, participants from white ethnic and more affluent socioeconomic backgrounds were overrepresented. Participation did not vary by age, sex, or clinical characteristics. | The poor recruitment rate from children and adolescents from ethnic minority backgrounds may be because that recruitment letters were sent out in English only.  Most participants were white in both studies 1 and 2, and in study 2, there was an underrepresentation of Asian participants. |
| Tan, L. L., & Denson, L. (2019). Bilingual and multilingual psychologists practising in Australia: an exploratory study of their skills, training needs and experiences. *Australian Psychologist*, *54*(1), 13-25.  (Tan and Denson, 2019) | Australia | **Type of study:**  Mixed method: Survey and telephone interviews  **Description:**  An online survey including demographic and practice information.  Supplementary telephone interviews with 11 of the participants. | N = 38 bilingual/multilingual psychologists working in Australia in 2015.  N = 11 participants undertook supplementary telephone interviews which along with survey responses were transcribed for qualitative thematic analysis. | No outcome measures were included in the survey. | Most participants trained in English. They expressed concerns about their application of psychological concepts in other languages, despite good conversational fluency. Participants highlighted language barriers to entering the profession; limited multicultural and multilingual training and supervision in Australia; and the need for more transcultural mental health resources, particularly for small/new migrant communities and people outside large cities. | The authors stated that the Psychology profession must actively support supervision, professional development, and practice in community languages. |
| Wang, L., & Kwak, M. J. (2015). Immigration, barriers to healthcare and transnational ties: A case study of South Korean immigrants in Toronto, Canada. *Social Science & Medicine*, *133*, 340-348.  (Wang and Kwak, 2015) | Canada | **Type of study:**  Mixed method study with a focus on qualitative data  **Description:** A  mixed-method study, with an emphasis on the qualitative facet of the study, to capture insights into the experience of Korean immigrants in seeking and receiving healthcare | N = 8 focus groups (n=54 in total; 81.5% female and 18.5% male). | No outcome measures were used in this qualitative study | Almost all the participants preferred to have a Korean-speaking family physician, while six out of 10 were able to find one. Language barriers, such as difficulty describing symptoms in English, lack of familiarity with medical terms, and difficulty understanding physicians' instructions in English, were experienced by both long term and new immigrants:  “I have lived here a long time so my English is okay for basic things. But when my symptoms are very complicated and complex it is true that my English is not sufficient. It is not the same as being a Canadian whose mother tongue is English. I cannot express completely my symptoms to the doctors. That is something I have experienced hundreds of times”. | There is some provision for South Korean immigrants in Canada (4 out of 10 people could access a Korean speaking general practitioner), however 6 of 10 could not. Therefore patients with children tended not to access healthcare in Canada.  Nearly half of the participants indicated that they did not receive timely care in Canada. The “long wait time” for diagnosis, treatment and operation was the main reason for seeking alternative resources in South Korea, where people can “get the service the day they arrive.”  This paper has highlighted that the screening process is not as solid as it could be, for example one senior patient was told by her healthcare professional:  “If you do not know the word, then it probably has no relevance for you… so just put ‘no’ on those.”  The findings from this paper highlights the importance of language concordance and training of healthcare professionals in dealing with patients from ethnic minority backgrounds. |
| Wang, A. M. Q., Yung, E. M., Nitti, N., Shakya, Y., Alamgir, A. K. M., & Lofters, A. K. (2019). Breast and colorectal cancer screening barriers among immigrants and refugees: a mixed-methods study at three community health centres in Toronto, Canada. *Journal of immigrant and minority health*, *21*(3), 473-482.  (Wang *et al.*, 2019) | Canada | **Type of study:**  A mixed-methods study  **Description:**  This mixed methods study involved a retrospective chart review of client data (quantitative data) and two focus groups (qualitative data) with Access Alliance primary care providers (PCPs). | The inclusion criteria were active clients aged 50–74 from all three Access Alliance Toronto sites, who were defined as having had at least one clinic encounter between November 1^st^, 2012 and October 31^st^ , 2015 inclusive.  Quantitative chart review: n = 2420 registered charts  Qualitative focus groups: n = 13 participants approached | No outcome measures were used in this mixed methods study. | Regarding language barriers, providers described the need for interpreters as being important but time and energy consuming. Additionally, participants noted that mammography forms and self-administered Faecal Occult Blood Test (FOBT) kits are only available in English or French.  “We have 30 minutes per appointment typically, but when they’re dealing with seven other issues and language line, interpretation services, you don’t necessarily have that much time to explain in detail the importance of screening” (page 477). | This moderate quality mixed method study of recent immigrants and refugees identifies barriers to breast and colorectal cancer screening and supports potential solutions including culturally-congruent peer workers (including language specific), targeted screening workshops, and language specific visual screening aids. Further work is needed to address the unique healthcare needs of immigrants and refugees in Canada. |

### Table A8 Research syntheses (n=18)

| **Citation** | **Country** | **Study details** | **Participants** | **Outcomes** | **Results** | **Overview** |
| --- | --- | --- | --- | --- | --- | --- |
| Beresford, P. (2007). User involvement, research and health inequalities: developing new directions. *Health & Social Care in the Community*, *15*(4), 306-312.  (Beresford, 2007) | England, UK | **Type of study:** A review  Description: A descriptive narrative. | No participants described as it was a descriptive narrative. | To develop new directions in health inequalities research. | There is a recognition of the importance of language in ensuring inclusion, and developing appropriate language policy and practice (Beresford & Croft 1993, Ward 1997, Morris 1998, Wilkinson 2002). | This was a descriptive narrative, not based on a systematic review or meta-analysis methodology.  The authors noted the importance of involvement of all groups of service users to get involved in research, whatever their circumstances or identity. |
| Gibbs, B. K., Nsiah-Jefferson, L., McHugh, M. D., Trivedi, A. N., & Prothrow-Stith, D. (2006). Reducing racial and ethnic health disparities: exploring an outcome-oriented agenda for research and policy. *Journal of Health Politics, Policy and Law*, *31*(1), 185-218.  (Gibbs *et al.*, 2006) | USA | **Type of study:**  A review of American policies.  **Description:** A review of American policies to reduce racial and ethnic health disparities in research in the USA. | Eleven reports were initially selected, however after the inclusion criteria was applied, only n=4 reports were analysed. | No outcome measures were used (as this was a review). | The authors make the following recommendations:  1.The federal government, with researchers, should set a national agenda around Racial and Ethnic Health Disparities (REHD) reduction that is outcome oriented and encourages additional work to (a) apply, reconstruct, and refine the disparity reduction profile.  2. Use the disparity reduction profile correlations, when available, not to replace the in-depth qualitative work that should determine what is effective in communities and at what levels, but to provide a larger picture of efforts that can be followed over time.  3. Use the Disparity Index to help legislators and other stakeholders make reasonable decisions based on need, persistence of problems, and new challenges.  4. Consider other uses of the deprivation index to measure health services such as immunizations, primary health care, cancer screening and management, and substance use as well as other programs such as Women, Infants, Children, Food Stamps, and Medicare. Currently, most of these measures do not meet our criteria for use for developing a state disparity index, but maybe at some point when racial/ethnic data are improved, this combination of measuring changes in effort and activities and racial and ethnic health disparities (REHD) can be helpful. | This article makes the case for an outcome-oriented approach to reducing racial and ethnic health disparities and provides a summary of lessons learned. All four reports listed four or more of the following barriers as obstacles to providing services: 1. lack of accurate data to measure and document progress, shortage of minority-targeted health programs, limited technical assistance available to improve the quality of health care professionals, inadequate funding or lack of funding priorities, cultural and language barriers, limitations in data collection, an increase in the number of languages spoken in the United States, unexpected and rapid demographic changes, and geographic isolation. |
| Goode, T. D., Carter-Pokras, O. D., Horner-Johnson, W., & Yee, S. (2014). Parallel tracks: Reflections on the need for collaborative health disparities research on race/ethnicity and disability. *Medical care*, *52*(10 0 3), S3.  (Goode *et al.*, 2014) | USA | **Type of study:** A commentary  **Description:** This commentary reflects on the history, foci, and current status of these two separate tracks of health disparities research. A historical lens offers valuable insight as to why very little of the existing literature focuses on the intersection of race, ethnicity, and disability. | No participants as this was a commentary. | This was a commentary and no outcome measures were used. | Both quantitative and qualitative studies are needed to understand the experiences of people “at the intersection” of disability and ethnic group, and determine if the barriers they face are multiplied because of their unique status. Further, there is a need to create and study health disparity interventions that are culturally and linguistically competent for the diverse population of people with disabilities. This will require approaches to research that acknowledge and measure myriad cultural differences among people with disabilities effectively, rather than simply using race and ethnicity as proxies for culture. Diversity includes factors beyond disability and race and ethnicity. | Research in both disability and ethnicity frequently fails to address the multiple cultural identities within population groups.  This commentary, which is of low quality in terms of evidence, ends with suggestions for future research that addresses meeting the cultural and language needs of those with disabilities from ethnic minority backgrounds in the USA, which is a diverse country. |
| Hunt, S. M., & Bhopal, R. (2004). Self report in clinical and epidemiological studies with non-English speakers: the challenge of language and culture. *Journal of Epidemiology & Community Health*, *58*(7), 618-622.  (Hunt and Bhopal, 2004) | Scotland, UK | **Type of study:** A commentary  **Description:** A methodological commentary regarding including non-English speakers in research. | No participants as this was a commentary on theory and methods. | This was a commentary and no outcome measures were used. | It is important to bear in mind that the content of a questionnaire reflects not only the language of the originating country but also the standards, expectations, values, and preoccupations of both the researchers and the lay people involved in the developmental procedures. | The authors noted that even when people belonging to another culture speak fluent English they do not necessarily share the beliefs and values of native English speakers.  Therefore forward and back translation is not enough by itself.  “Translation into appropriate languages and back translation are necessary but insufficient steps” (P.619). |
| Jacobs, E., Chen, A. H., Karliner, L. S., Agger‐Gupta, N., & Mutha, S. (2006). The need for more research on language barriers in health care: a proposed research agenda. *The Milbank Quarterly*, *84*(1), 111-133.  (Jacobs *et al.*, 2006) | USA | **Type of study:** A review  **Description:** The authors found the two articles examined for their review through two systematic and thorough reviews of the literature (Jacobs et al. 2003; Karliner et al. 2005). Both reviews identified peer-reviewed journal articles through a systematic search of PubMed, PsychINFO, and Sociological Abstracts databases completed in 2003. | Two systematic reviews of the literature | This was a review and no outcome measures were used. | Three broad areas needing more research were discussed: 1) the ways in which language barriers affect health and health care  2) the efficacy of linguistic access service interventions  3) the costs of language barriers and efforts to overcome them. | Given the rapid increase in the number of Americans reporting that they speak English “less than very well,” there is a critical need for the research community to provide health care providers and policymakers with the evidence they require to design and effectively implement linguistically accessible services to limited English proficiency (LEP) patients.  The authors highlight aspects of costs, lost work time due to delayed diagnoses, unnecessary repeat visits, and preventable medication errors stemming from miscommunication in the medical encounter as factors to be considered when reducing language barriers in health care.  The authors note that:  “researchers need to explore the direct and indirect costs of language barriers for LEP patients and communities” (P.123). |
| Kaplan, J. B. (2014). The quality of data on “race” and “ethnicity”: Implications for health researchers, policy makers, and practitioners. *Race and Social Problems*, *6*(3), 214-236.  (Kaplan, 2014) | USA | **Type of study:** A commentary  **Description:** A commentary on the quality of data on “race” and “ethnicity” in the USA. | No participants as this was a commentary. | This was a commentary and no outcome measures were used. | This article outlines a series of issues that challenge assumptions about the quality of race/ethnicity data. While race/ethnicity classifications can approximate socially constructed identities for some groups of people under some circumstances, these classifications are inherently too imprecise to allow meaningful statements to be made about underlying biological or genetic differences between groups. | The authors highlight the fact that groups such as ‘latinos’ are not homogenous. There is no one clear definition of who belongs to a certain group and this can lead to adverse health consequences. |
| Lau, A. S., Chang, D. F., & Okazaki, S. (2010). Methodological challenges in treatment outcome research with ethnic minorities. *Cultural Diversity and Ethnic Minority Psychology*, *16*(4), 573.  (Lau, Chang and Okazaki, 2010) | USA | **Type of study:** A commentary  **Description:** Commentary discussing randomised controlled trials looking at specific challenges facing investigators conducting ethnically inclusive trials. | No participants as this was a commentary of RCT trials. | This was a commentary and no outcome measures were used. | It is imperative that trials provide assessment and treatment in the appropriate language(s) and dialect(s) spoken by the ethnic group under study. However, a meta-analytic review of 76 studies evaluating interventions culturally adapted for ethnic minorities revealed that 40% of the studies included only native English speakers. Of those trials including non-native English speakers, 25% provided treatment only in English. These numbers reflect the sheer difficulty of conducting RCT studies with non-native English speakers.  Treatment as usual in some community settings for ethnic minorities may be no treatment or substandard treatment due to multiple barriers that prevent access to even minimally effective care (e.g., lack of language-matched providers). | Randomised controlled trials generally do not cater for those whose primary language is not English.  Treatments are provided in English mostly, and very rarely in the preferred language of the person with limited English proficiency.  Results of randomised controlled trials may well be more robust if there was better concordance between the patient’s language and the language of the intervention. |
| Martinez, I. L., Carter-Pokras, O., & Brown, P. B. (2009). Addressing the challenges of Latino health research: participatory approaches in an emergent urban community. *Journal of the National Medical Association*, *101*(9), 908-914.  (Martinez, Carter-Pokras and Brown, 2009) | USA | **Type of study:** A review  **Description:** The authors described review steps taken and describe lessons learned in using a participatory approach to broadly assess and address the health of urban-dwelling Latinos in Baltimore, Maryland, through the adaptation of Community Based Participatory Research (CBPR) principles. | Interviews with ex-nurses no longer working in the health field. | This was a review and no outcome measures were used. | While Spanish is a unifying language among most Latin American immigrants, dialects vary and, in some cases, other language minorities exist. The authors found that while Spanish-language fluency facilitates communication and contributes to a sense of familiarity and comfort, fluency or Latin American heritage does not guarantee community acceptance or trust. It may be more important to convey respect for community concerns and values through empathic listening, acknowledgement and understanding of community issues, and honouring of commitments. | Health research and health care access for Latinos in Baltimore (and elsewhere) present particular challenges, beyond the language and cultural barriers that might be expected. |
| Morville, A. L., & Erlandsson, L. K. (2016). Methodological challenges when doing research that includes ethnic minorities: a scoping review. *Scandinavian journal of occupational therapy*, *23*(6), 405-415.  (Morville and Erlandsson, 2016) | Sweden | **Type of study:** Scoping review  **Description:** Scoping review of methodological challenges when doing research that includes ethnic minorities. | **Number of articles included:** 21  The literature came mainly not only from the American continent, but also from the Netherlands and Sweden. | This was a scoping review and no outcome measures were used. | Five themes were identified including the following of relevance to this systematic review:  **Defining and recruiting samples:** Not many papers defined ‘ethnic minorities’. This influences sampling and recruitment into the research studies.  **Lack of appropriate instruments:** Very few instruments and assessments have had their psychometric properties tested in regard to the use of language and the understanding of the concepts.  **Data collection:** Using an interpreter during data collection gives some of the same problems as the translation of questionnaires. Using a verbatim translation during an interview will not make sense in most languages; and as with the questionnaires, the interpreting should capture the meaning of the interviewer’s question. However, the problem of using this approach might give the interpreter the responsibility of taking on a leading role, explaining the meaning of concepts and questions, and thus putting several roles upon the interpreter, and a large margin for errors | The results showed methodological issues concerning the entire research process from defining and recruiting samples, the conceptual understanding, lack of appropriate instruments, data collection using interpreters to analysing data.  In terms of language issues, Weitzman and Levkoff [45] and Small et al. [46] describe the option of using bilingual interviewers from the same ethnic minority, but as with interpreters the results are dependent on the interviewers’ style and cultural knowledge of both cultures. (P.412). |
| Murray, S., & Buller, A. M. (2007). Exclusion on grounds of language ability–a reporting gap in health services research?. *Journal of health services research & policy*, *12*(4), 205-208.  (Murray and Buller, 2007) | England, UK | **Type of study:** A review  **Description:** Retrospective review of 207 ‘original research pieces’ published in the BMJ. Inclusion criteria: articles published in 2003 and 2004 reporting research entailing direct communication between a researcher and health service users. | No participants as this was a review. 207 ‘original research pieces’ published in the BMJ. | No outcome measures were used in this retrospective review | Eighty-four percent of the research articles did not engage with language issues at all. For most papers it was impossible to ascertain whether research was carried out in a monolingual population or whether researchers had electively excluded non-primary language speakers in their recruitment procedures. Over half (n = 34) of the papers that mentioned language did so in relation to exclusion criteria, usually without further comment. | The majority of the papers included in the review did not mention language matters. The issues raised in this review from 2007 are also still relevant in 2021.  Members of ethnic minority language groups are still underrepresented in English only research.  The authors noted: “This seems to be a considerable deficiency in current research reporting practice” (P.206).  Checklist for the reporting of language issues relating to the recruitment of study participants:  1 Is the language composition of the population to be studied/ from which the sample is to be drawn clearly described? 2 Are the eligibility criteria for the sample clearly defined in relation to the language(s) of the population? 3 When potential participants are excluded based on language: Is this exclusion made explicit in the exclusion criteria? Is this exclusion justified? Is there any comment made concerning the implications for generalizability of findings resulting from this exclusion? 4 When non-primary language participants are included in a sample: Is there a sufficiently detailed description on the language related procedures followed to obtain informed consent, and the rationale behind these? |
| Nishita, C., & Browne, C. (2013). Advancing research in transitional care: Challenges of culture, language and health literacy in Asian American and native Hawaiian elders. *Journal of health care for the poor and underserved*, *24*(1), 404-418.  (Nishita and Browne, 2013) | Hawaii, USA | **Type of study:** A review  **Description:**  The literature review included a broad review of long-term care policy directions that support transitions from institutional settings, and evidence based models in transitional care. | No participants as this was a broad literature review looking at policy directions that support transitions in care. | No outcome measures were used in this literature review. | The paper presents a conceptual framework and proposes a six-point research agenda that includes family assessments of health literacy abilities, exploring the relationship between culture, health, and decision-making, and the development/adaptation of transitional care planning tools. | Health clinicians and other professionals must understand the influence of culture and language on health literacy and preferences for care, and residents and their families must be health literate so that they are able to make informed decisions in long-term transitional care. |
| Premji, S., Kosny, A., Yanar, B., & Begum, M. (2020). Tool for the meaningful consideration of language barriers in qualitative health research. *Qualitative health research*, *30*(2), 167-181.  (Premji *et al.*, 2020) | Canada | **Type of study:**  A review  **Description:** Review of authors own papers on the topic of language barriers in qualitative health research. | **Number of included studies:** 6 studies by the author were included.  There were between 25 and 110 participants in each of the included papers. | No outcome measures used in this broad literature review. | There remain gaps and debates with respect to the relevant ethical and methodological guidance set forth by funding agencies and institutions and proposed in the scientific literature. This article adds to knowledge in this area by contributing the experiences, observations, and recommendations of the authors, including around the issue of conducting research in contexts of more or less linguistic diversity. | The authors experiences may assist other health researchers conducting qualitative interviews in languages other than English.  In Canada, the Tri-Council Policy Statement or TCPS 2 (2014) on ethical conduct for research involving humans notes that researchers shall not exclude individuals from the opportunity to participate in research on the basis of attributes such as culture, language, religion, race, disability, sexual orientation, ethnicity, linguistic proficiency, gender or age, unless there is a valid reason for the exclusion. (article 4.1) |
| Rose, A. L., & Cheung, M. (2012). DSM-5 research: assessing the mental health needs of older adults from diverse ethnic backgrounds. *Journal of Ethnic and Cultural Diversity in Social Work*, *21*(2), 144-167.  (Rose and Cheung, 2012) | USA | **Type of study:**  A review  **Description:** An analysis of 54 articles published between 2001 and 2011 in 4 social science databases | No participants , but 54 articles published between 2001 and 2011 in 4 social science databases | The main focus was on the DSM-5 mental health classification system. | Many articles used in this review discuss the inadequacies of the DSM usage, which limited the diagnoses of people of colour and further increased mistrust and disparities. If a person is unable to receive a proper diagnosis of a mental health condition, it is assumed that proper treatment will never be reached | The authors recommend that changes are made to the DSM-5 which would make it easier to diagnose and classify mental disorders related to ‘people of colour’.  Language barriers sometimes stand in the way of elderly people who are not first language English in seeking a diagnosis of a mental health condition. |
| Schwei, R. J., Del Pozo, S., Agger-Gupta, N., Alvarado-Little, W., Bagchi, A., Chen, A. H., ... & Jacobs, E. A. (2016). Changes in research on language barriers in health care since 2003: A cross-sectional review study. *International journal of nursing studies*, *54*, 36-44.  (Rebecca J Schwei *et al.*, 2016) | USA and Canada | **Type of study:**  A review  **Description:** A cross-sectional review of research literature on language barriers in health care | 136 studies prior to 2003 and 426 studies from 2003 to 2010 were included in the review. | No outcome measures were used in this review. | The authors suggest that researchers worldwide should move away from simply documenting the existence of language barriers and should begin to focus their research on documenting how language concordant care influences patient outcomes, providing evidence for interventions that mitigate language barriers, and evaluating the cost effectiveness of providing language concordant care to patients with language barriers | There is enough evidence of language barriers and therefore the authors suggest that future research should concentrate on the effectiveness and cost-effectiveness of providing language concordant care. |
| Sue, S., & Dhindsa, M. K. (2006). Ethnic and racial health disparities research: Issues and problems. *Health Education & Behavior*, *33*(4), 459-469.  (Sue and Dhindsa, 2006) | USA | **Type of study:**  A review  **Description:**  A review addressing ethnic and racial health and health care disparities research. | No participants as this was a review. | No outcome measures were used in this review | The authors make five recommendations to address ethnic and racial health and health care disparities.  The third recommendation was about language:  Recommendation 3:  Studying different ethnic and racial groups, measures used to assess symptoms, illness, health practices, cultural attitudes, and so on need to be cross-culturally validated. Otherwise, ethnic comparisons can yield misleading information disparities. | The subtlety of culture needs to be recognised as an important factor in health research, and also, ethnic groups should not be clusters without clear explanation. |
| Waheed, W., Hughes-Morley, A., Woodham, A., Allen, G., & Bower, P. (2015). Overcoming barriers to recruiting ethnic minorities to mental health research: a typology of recruitment strategies. *BMC psychiatry*, *15*(1), 1-11.  (Waheed *et al.*, 2015) | England, UK | **Type of study:**  A review  **Description:**  The authors extracted data from a previous systematic review on the strategies used to overcome recruitment barriers | 9 papers were included in the original systematic review. | No outcome measures were included as part of this review. | The typology of strategies to overcome barriers provided by the authors, provides guidance on achieving higher rates of recruitment. However, the research to quantify the positive impact of these strategies on recruitment has not been conducted. | The authors suggest that bilingual and bicultural staff should be utilised in research. This helps in better communication in participant’s preferred language and provides greater sensitivity when selecting language and common expressions.  Having bilingual staff was seen as ‘critically important’.  Careful translation of research tools was also seen as important as well as high quality cultural competency training for research staff.  Cultural sensitivity is especially important when approaching women from ethnic minorities. |
| Wells, A. A., & Zebrack, B. (2007). Psychosocial barriers contributing to the under-representation of racial/ethnic minorities in cancer clinical trials. *Social Work in Health Care*, *46*(2), 1-14.  (Wells and Zebrack, 2007) | USA | **Type of study:**  A review  **Description:** Utilizing a social-ecological perspective, with particular emphasis on socio-cultural aspects, this paper presents a multi-level categorization of psychosocial barriers to participation in oncology clinical trials among racial and ethnic minority groups. | No participants in this review. Studies were not selected | No outcome measures were described in this review. | It is especially important to emphasize important elements of cultural relevance because many of the barriers to participation occur within a socio-cultural context that impacts one’s beliefs and attitudes. In addition to obvious language barriers which interfere with the patients’ understanding of information, socio-cultural barriers include community mistrust and scepticism related to past historical injustices in research. | Socio-cultural barriers include other issues, alongside language. This may include mistrust of researchers and the research process. |
| Yildiz, C., & Bartlett, A. (2011). Language, foreign nationality and ethnicity in an English prison: implications for the quality of health and social research. *Journal of Medical Ethics*, *37*(10), 637-640.  (Yildiz and Bartlett, 2011) | England, UK | **Type of study:**  A review  **Description:**  **Review of** studies to understand how Foreign National prisoners were included or excluded from research undertaken in a women’s prison in London. | N = 27 studies were included | No outcome measures were used in this review. | One of the 27 papers stated that they translated research information forms into two other languages apart from English and then excluded data from women with limited English as the women did not speak those particular two languages other than English. | Only 4 out of 27 papers mentioned translation into other languages other than English despite Foreign National prisoners constituting 33% of the total population of the prison at the time of the study in July 2010. |

### Table A9 Case report (n=1)

| **Citation** | **Country** | **Study details** | **Participants** | **Outcomes** | **Results** | **Overview** |
| --- | --- | --- | --- | --- | --- | --- |
| Du, F. H. (2018). Gray Areas in Language-Concordant Healthcare: a Graduating Medical Student’s Reflection on the Experience and Research on Language and Cultural Competence. *Journal of Cancer Education*, *33*(2), 493-496.  (Du, 2018) | USA | **Type of study:**  Case study  **Description:** Reflection of graduate medical student. Real-life example of different healthcare examples regarding language and communication. | No participants but reflection of a recent real-life healthcare example of the importance of language and communication. | No outcome measures utilized in this reflection. | The author noted that language concordant care providers and professional medical interpreters are invaluable for limited English proficiency families. Healthcare teams can also come together to create inclusive environments that address language barriers and beyond for limited English proficiency patients.  Author believed that physicians with some proficiency in another language (e.g., Spanish) tended to overestimate their own ability. | Language barriers are not the only barriers that limited English proficiency persons experience in healthcare.  Of note, having language-concordant providers is of even greater importance to LEP patients who also have limited education compared to those with more education, indicating a greater need to a group more vulnerable to health disparities  Regardless of a clinician’s language proficiency, tailoring care and connecting with limited English proficiency patients and families by any means during times of illness can be comforting and appreciated. |
