## Supplementary material for "A systematic review of the experiences of minority language users in health and social care research"

### Supplementary Material 1 – Quality appraisal checklists

Quality appraisal tables are presented below for included studies according to the methodology used for the study, these include 5 cross sectional studies, 7 systematic reviews, 2 meta-analyses, 22 qualitative studies, 9 mixed methods studies, 19 were research syntheses studies, and 1 case report study (n=74 studies included in total).

Five cross-sectional studies were quality appraised (see Table A1)

#### Table A1 Cross-sectional studies – quality appraisal checklist (Moola et al., 2020).

| **Citation** | **Q1. Were the criteria for inclusion in the sample clearly defined?** | **Q2. Were the study subjects and the setting described in detail?** | **Q3. Was the exposure measured in a valid and reliable way?** | **Q4. Were objective, standard criteria used for measurement of the condition?** | **Q5. Were confounding factors identified?** | **Q6. Were strategies to deal with confounding factors stated?** | **Q7. Were the outcomes measured in a valid and reliable way?** | **Q8. Was appropriate statistical analysis used?** |
| --- | --- | --- | --- | --- | --- | --- | --- | --- |
| Durbin et al (2017) | Yes | Yes | N/A | Yes | Yes | No | Yes | Yes |
| Kale & Syed. (2010) | Yes | Yes | N/A | Yes | Yes | Yes | Yes | Yes |
| Peek et al (2012) | Yes | Yes | N/A | Yes | Yes | Unclear | Yes | Yes |
| Silveira et al (2020) | Yes | Yes | N/A | Yes | Yes | Yes | Yes | Yes |
| Wilk et al (2018) | Yes | Yes | N/A | No | Yes | No | No | Unclear |

Seven systematic reviews were quality appraised (see Table A2).

#### Table A2 Systematic Reviews - quality appraisal checklist (Aromataris et al., 2015)

| **Citation** | **Q1. Is the review question clearly and explicitly stated?** | **Q2. Were the inclusion criteria appropriate for the review question?** | **Q3. Was the search strategy appropriate?** | **Q4. Were the sources and resources used to search for studies adequate?** | **Q5. Were the criteria for appraising studies appropriate?** | **Q6. Was critical appraisal conducted by two or more reviewers independently?** | **Q7. Were there methods to minimize errors in data extraction?** | **Q8. Were the methods used to combine studies appropriate?** | **Q9. Was the likelihood of publication bias assessed?** | **Q10. Were recommendations for policy and/or practice supported by the reported data?** | **Q11. Were the specific directives for new research appropriate?** |
| --- | --- | --- | --- | --- | --- | --- | --- | --- | --- | --- | --- |
| Brown et al (2014) | No | Unclear | Yes | Yes | No | Yes | Yes | Yes | Yes | Yes | Yes |
| Castillo et al (2019) | No | Yes | Yes | Yes | Unclear | Yes | Yes | Yes | Yes | Yes | Yes |
| Chowdhury et al (2021) | No | Yes | Yes | Yes | Yes | Yes | No | Yes | No | Yes | Yes |
| Di Pietro & Illes (2014) | No | Yes | Yes | Yes | No | No | Unclear | Unclear | No | Yes | Yes |
| Huang et al (2019) | No | Yes | Yes | Yes | Yes | Yes | Yes | Unclear | Yes | Yes | Yes |
| Joo, & Liu (2020) | No | Yes | Yes | Yes | Yes | Yes | Yes | Unclear | No | No | Yes |
| Woodall et al (2010) | Yes | Yes | Yes | Yes | No | No | Unclear | Unclear | No | Yes | Yes |

Two meta analyses were data appraised (see Table A3)

#### Table A3 Meta analyses - quality appraisal checklist (Aromataris et al., 2015)

| **Citation** | **Q1. Is the review question clearly and explicitly stated?** | **Q2. Were the inclusion criteria appropriate for the review question?** | **Q3. Was the search strategy appropriate?** | **Q4. Were the sources and resources used to search for studies adequate?** | **Q5. Were the criteria for appraising studies appropriate?** | **Q6. Was critical appraisal conducted by two or more reviewers independently?** | **Q7. Were there methods to minimize errors in data extraction?** | **Q8. Were the methods used to combine studies appropriate?** | **Q9. Was the likelihood of publication bias assessed?** | **Q10. Were recommendations for policy and/or practice supported by the reported data?** | **Q11. Were the specific directives for new research appropriate?** |
| --- | --- | --- | --- | --- | --- | --- | --- | --- | --- | --- | --- |
| Angus et al (2012) | Yes | Yes | Yes | Yes | Yes | Yes | Yes | Yes | No | Yes | Yes |
| Clarke et al (2013) | Yes | Unclear | Yes | Unclear | Yes | Yes | Yes | Unclear | Yes | Yes | Yes |

Twenty two qualitative studies were quality appraised (see Table A4).

#### Table A4 Qualitative studies (Lockwood, Munn, & Porritt, 2015)

| **Citation** | **Q1 Is there congruity between the stated philosophical perspective and the research methodology?** | **Q2 Is there congruity between the research methodology and the research question or objectives?** | **Q3 Is there congruity between the research methodology and the methods used to collect data?** | **Q4 Is there congruity between the research methodology and the representation and analysis of data?** | **Q5 Is there congruity between the research methodology and the interpretation of results?** | **Q6 Is there a statement locating the researcher culturally or theoretically?** | **Q7 Is the influence of the researcher on the research, and vice- versa, addressed?** | **Q8 Are participants, and their voices, adequately represented?** | **Q9 Is the research ethical according to current criteria or, for recent studies, and is there evidence of ethical approval by an appropriate body?** | **Q10 Do the conclusions drawn in the research report flow from the analysis, or interpretation, of the data?** |
| --- | --- | --- | --- | --- | --- | --- | --- | --- | --- | --- |
| Akhavan and Karlesen (2013) | Yes | Yes | Yes | Yes | Yes | Unclear | Unclear | Yes | Yes | Yes |
| Betancourt et al (2015) | Yes | Yes | Yes | Yes | Yes | Yes | Yes | Yes | No | Yes |
| Claydon-Platt et al (2014) | Yes | Yes | Yes | Yes | Yes | Unclear | No | Yes | Yes | Yes |
| De La Torre (2010) (Doctoral thesis) | Yes | Yes | Yes | Yes | Yes | Yes | Yes | Yes | Yes | Yes |
| Dingoyan et al (2012) | Yes | Yes | Yes | Yes | Yes | Yes | No | Yes | No | Yes |
| Doyle (2016) | Yes | Yes | Yes | Yes | Yes | Yes | Yes | Yes | No | Yes |
| Fisher (2011) | Yes | Yes | Yes | No | No | Yes | Yes | No | Yes | No |
| French and Stravropoulou (2016) | Yes | Yes | Yes | Yes | Yes | Yes | Yes | Yes | Yes | Yes |
| Gaston-Johansson et al (2008) | Yes | Yes | Yes | Yes | Yes | No | No | Yes | No | Yes |
| Haley et al (2017) | Yes | Yes | Yes | Yes | Yes | Unclear | No | Yes | Yes | Yes |
| Hunter-Adams & Rother (2017) | Yes | Yes | Yes | Yes | Yes | No | No | Yes | Yes | Yes |
| Johnsen et al (2020) | Yes | Yes | Yes | Yes | Yes | Yes | No | Yes | Yes | Yes |
| MacFarlane (2009) | Yes | Yes | Yes | Yes | Yes | Yes | Yes | Yes | Yes | Yes |
| O’Connor et al (2018) | Yes | Yes | Yes | Yes | Yes | Yes | Yes | Yes | No | Yes |
| Robinson and Trochim (2007) | Yes | Yes | Yes | Yes | Yes | Unclear | Unclear | Yes | No | Yes |
| Sadavoy et al (2004) | Yes | Yes | Yes | Unclear | Unclear | No | No | Yes | No | Yes |
| Schildmann et al (2016) | Yes | Yes | Yes | Yes | Yes | Unclear | No | Yes | Yes | Yes |
| Shattell et al (2008) | Yes | Yes | Yes | Yes | Yes | Unclear | No | Yes | No | Yes |
| Squires et al (2019) | Yes | Yes | Yes | Yes | Yes | Unclear | No | Yes | Yes | Yes |
| Strohschein et al (2010) | Yes | Yes | Yes | Yes | Yes | Yes | Yes | Yes | Yes | Unclear |
| Tatari et al (2020) | Yes | Yes | Yes | Yes | Yes | Yes | Yes | Yes | Yes | Yes |
| Vandan et al (2020) | Yes | Yes | Yes | Yes | Yes | Yes | No | Yes | Yes | Yes |

Ten quantitative studies were critically appraised. One was a cohort study and the other nine were survey studies (see Tables A5 and A6).

#### Table A5 Cohort Studies (Moola et al., 2020)

| Citation | Q1. Were the two groups similar and recruited from the same population? | Q2. Were the exposures measured similarly to assign people to both exposed and unexposed groups? | Q3. Was the exposure measured in a valid and reliable way? | Q4. Were confounding factors identified? | Q5. Were strategies to deal with confounding factors stated? | Q6. Were the groups/ participants free of the outcome at the start of the study (or at the moment of exposure)? | Q7. Were the outcomes measured in a valid and reliable way? | Q8. Was the follow up time reported and sufficient to be long enough for outcomes to occur? | Q9. Was follow up complete, and if not, were the reasons to loss to follow up described and explored? | Q10. Were strategies to address incomplete follow up utilized? | Q11. Was appropriate statistical analysis used? |
| --- | --- | --- | --- | --- | --- | --- | --- | --- | --- | --- | --- |
| Blanchet et al (2017) | Yes | Yes | Yes | Yes | Yes | Yes | Yes | Yes | No | No | Yes |

Nine survey studies were included and critically appraised (Center for Evidence Based Management, 2005)

#### Table A6 Survey studies (Center for Evidence Based Management, 2005)

| Citation | Q1 Did the study address a clearly focused question / issue? | Q2 Is the research method (study design) appropriate for answering the research question? | Q3 Is the method of selection of the subjects (employees, teams, divisions, organizations) clearly described? | Q4 Could the way the sample was obtained introduce (selection) bias? | Q5 Was the sample of subjects representative with regard to the population to which the findings will be referred? | Q6 Was the sample size based on pre-study considerations of statistical power? | Q7 Was a satisfactory response rate achieved? | Q8 Are the measurements (questionnaires) likely to be valid and reliable? | Q9 Was the statistical significance assessed? | Q10 Are confidence intervals given for the main results? | Q11 Could there be confounding factors that haven’t been accounted for? | Q12. Can the results be applied to your organization? |
| --- | --- | --- | --- | --- | --- | --- | --- | --- | --- | --- | --- | --- |
| Brisset et al (2014) | Yes | Yes | Yes | No | Yes | No | Yes | No | Yes | N/A | No | N/A |
| Carlini et al (2015) | Yes | Yes | Yes | No | Yes | No | Yes | No | Yes | No | No | N/A |
| Coustasse et al (2009) | Yes | Yes | Yes | No | Yes | No | Yes | No | Yes | Yes | No | N/A |
| Falla et al (2017) | Yes | Yes | Yes | No | Yes | No | Yes | No | No | No | No | N/A |
| Greene et al (2013) | Yes | Yes | Yes | No | Yes | No | No | No | No | No | No | N/A |
| Hahm et al (2008) | Yes | Yes | Yes | No | Yes | Yes | Yes | No | Yes | Yes | No | N/A |
| Kim et al (2011) | Yes | Yes | Yes | No | Yes | Yes | Unclear | No | Yes | Yes | No | N/A |
| Lu et al (2014) | Yes | Yes | Yes | No | Yes | No | Unclear | No | Yes | N/A | No | N/A |
| Mosconi et al (2016) | Yes | Yes | Yes | No | Yes | No | No | No | No | N/A | No | N/A |

Nine mixed method studies were critically appraised, see Table A6.

#### Table A7 Mixed methods studies (Hong et al., 2018)

| **Category of study design** | **Methodological quality criteria** | **Ahlmark et al 2015** | **Bhuiyan et al (2019)** | **Chirewa (2012)** | **Eriksson-Sjoo et al (2012)** | **Mowlabaccus & Jodheea-Jutton (2020)** | **Tadic et al (2010)** | **Tan and Denson (2019)** | **Wang and Kwak (2015)** | **Wang et al (2019)** |
| --- | --- | --- | --- | --- | --- | --- | --- | --- | --- | --- |
| **Screening questions** | S1. Are there clear research questions? | Yes | Yes | No | Yes | Yes | Yes | Unclear | Unclear | Yes |
|  | S2. Do the collected data allow to address the research questions? | Yes | Yes | Yes | Yes | Yes | Yes | Yes | Yes | Yes |
| **1. Qualitative** | 1.1. Is the qualitative approach appropriate to answer the research question? | Yes | Yes | Yes | Yes | Yes | Unclear | Yes | Yes | Yes |
|  | 1.2. Are the qualitative data collection methods adequate to address the research question? | Yes | Yes | Yes | Yes | Yes | Unclear | Yes | Yes | Yes |
|  | 1.3. Are the findings adequately derived from the data? | Yes | Yes | Unclear | Yes | Yes | No | Yes | Yes | Yes |
|  | 1.4. Is the interpretation of results sufficiently substantiated by data? | Yes | Unclear | No | Yes | Yes | No | Yes | Yes | Yes |
|  | 1.5. Is there coherence between qualitative data sources, collection, analysis and interpretation? | Yes | Yes | Unclear | Yes | Yes | No | Yes | Yes | Yes |
| **2. Quantitative randomized controlled trials** | 2.1. Is randomization appropriately performed? | N/A | N/A | N/A | N/A | N/A | N/A | N/A | N/A | N/A |
|  | 2.2. Are the group comparable at baseline? | N/A | N/A | N/A | N/A | N/A | N/A | N/A | N/A | N/A |
|  | 2.3. Are there complete outcome data? | N/A | N/A | N/A | N/A | N/A | N/A | N/A | N/A | N/A |
|  | 2.4. Are outcome assessors blinded to the intervention provided? | N/A | N/A | N/A | N/A | N/A | N/A | N/A | N/A | N/A |
|  | 2.5. Did the participants adhere to the assigned intervention? | N/A | N/A | N/A | N/A | N/A | N/A | N/A | N/A | N/A |
| **3. Quantitative non-randomized** | 3.1. Are the participants representative of the target population? | Yes | Unclear | No | Yes | Yes | No | N/A | Yes | N/A |
|  | 3.2. Are measurements appropriate regarding both the outcome and the intervention (or exposure)? | Yes | Unclear | Unclear | Yes | Yes | Yes | N/A | N/A | N/A |
|  | 3.3. Are there complete outcome data? | Yes | Unclear | No | Yes | Yes | Yes | N/A | N/A | N/A |
|  | 3.4. Are the confounders accounted for in the design and analysis? | Yes | Unclear | No | Unclear | Yes | Unclear | N/A | Yes | N/A |
|  | 3.5. During the study period, is the intervention administered (or exposure) as intended? | N/A | Unclear | N/A | N/A | N/A | N/A | N/A | N/A | N/A |
| **4. Quantitative descriptive** | 4.1. Is the sampling strategy relevant to address the research question? | Yes | Yes | No | Yes | Yes | Yes | Yes | Yes | Unclear |
|  | 4.2. Is the sample representative of the target population? | Yes | Unclear | No | Yes | Yes | No | Yes | Yes | No |
|  | 4.3. Are the measurements appropriate? | Yes | Unclear | Unclear | Yes | Yes | Yes | Yes | Yes | Yes |
|  | 4.4. Is the risk of nonresponse bias low? | Yes | Unclear | No | No | Yes | No | Yes | Unclear | Yes |
|  | 4.5. Is the statistical analysis appropriate to answer the research question? | Yes | Yes | No | Yes | Yes | Yes | N/A | Yes | Yes |
| **5. Mixed methods** | 5.1. Is there an adequate rationale for using a mixed methods design to address the research question? | Yes | No | Yes | Unclear | Yes | No | Unclear | Yes | No |
|  | 5.2. Are the different components of the study effectively integrated to answer the research question? | Yes | Unclear | No | Yes | Yes | No | Yes | Unclear | No |
|  | 5.3. Are the outputs of the integration of qualitative and quantitative components adequately addressed? | Yes | No | Yes | Yes | Yes | No | Yes | Yes | Yes |
|  | 5.4. Are divergences and inconsistencies between quantitative and qualitative results adequately addressed? | Yes | No | Unclear | Yes | Yes | No | Yes | Yes | No |
|  | 5.5. Do the different components of the study adhere to the quality criteria of each tradition of the methods involved? | Yes | Unclear | No | Yes | Yes | No | Yes | Yes | Yes |

Nineteen research syntheses studies were quality appraised (see Table A8).

#### Table A8 Research syntheses (Aromataris et al., 2015)

| **Citation** | **Q1. Is the review question clearly and explicitly stated?** | **Q2. Were the inclusion criteria appropriate for the review question?** | **Q3. Was the search strategy appropriate?** | **Q4. Were the sources and resources used to search for studies adequate?** | **Q5. Were the criteria for appraising studies appropriate?** | **Q6. Was critical appraisal conducted by two or more reviewers independently?** | **Q7. Were there methods to minimize errors in data extraction?** | **Q8. Were the methods used to combine studies appropriate?** | **Q9. Was the likelihood of publication bias assessed?** | **Q10. Were recommendations for policy and/or practice supported by the reported data?** | **Q11. Were the specific directives for new research appropriate?** |
| --- | --- | --- | --- | --- | --- | --- | --- | --- | --- | --- | --- |
| Beresford (2007) | No | No | No | No | No | No | No | Yes | No | Yes | Yes |
| Brown et al (2014) | Yes | Yes | Yes | Yes | No | No | No | Yes | No | Yes | Yes |
| Di Petro and Illes (2014) | Yes | Yes | Yes | Yes | No | No | No | Yes | No | Yes | Yes |
| Gibbs et al (2006) | Yes | Yes | Yes | Yes | No | No | No | Yes | N/A | Yes | Yes |
| Goode et al (2014) | No | N/A | N/A | No | No | No | No | No | No | Yes | Yes |
| Hunt and Bhopal (2003) | Unclear | N/A | N/A | No | No | No | No | No | No | Yes | Yes |
| Jacobs (2006) | Yes | Yes | Yes | Yes | No | Unclear | No | Yes | No | Yes | Yes |
| Kaplan (2014) | No | N/A | N/A | Yes | No | No | No | Yes | No | Yes | Yes |
| Lau et al (2010) | No | N/A | N/A | No | No | No | No | Yes | No | Yes | Yes |
| Martinez et al (2009) | No | N/A | N/A | No | No | No | No | No | No | No | No |
| Morville & Erlandsson (2016) | Yes | Yes | Unclear | Yes | No | No | No | Yes | No | Yes | Yes |
| Murray and Buller (2007) | Yes | Yes | Yes | No | Unclear | Unclear | Unclear | Unclear | No | Yes | No |
| Nishita and Browne (2013) | Unclear | N/A | N/A | No | No | No | No | Yes | No | Yes | Yes |
| Premji et al (2020) | Yes | Yes | N/A | N/A | No | No | No | Yes | No | Yes | Yes |
| Rose and Cheung (2012) | Yes | Unclear | Yes | Yes | No | Yes | No | Unclear | No | Yes | Unclear |
| Schwei et al (2016) | Yes | Yes | Yes | Yes | No | No | No | Yes | No | Yes | Yes |
| Sue and Dhindsa | Yes | Yes | No | No | No | No | No | Yes | No | Yes | Yes |
| Waheed et al (2015)* | Yes | Yes | Yes | Unclear | No | No | No | Yes | No | Yes | Yes |
| Yildiz and Bartlett (2011) | Yes | Yes | Yes | Yes | No | No | No | Yes | No | Yes | Yes |

*The full systematic review is detailed in Brown et al (2014), listed in the Systematic Review table. Waheed et al (2015) is an extension to that systematic review.

One of the included papers was quality appraised as a case report (see Table A9).

#### Table A9 Case report - quality appraisal checklist (Munn et al., 2021)

| **Paper** | **Q1 Were patient’s demographic characteristics clearly described?** | **Q2 Was the patient’s history clearly described and presented as a timeline?** | **Q3 Was the current clinical condition of the patient on presentation clearly described?** | **Q4 Were diagnostic tests or assessment methods and the results clearly described?** | **Q5 Was the intervention(s) or treatment procedure(s) clearly described?** | **Q6 Was the post-intervention clinical condition clearly described?** | **Q7 Were adverse events (harms) or unanticipated events identified and described?** | **Q8 Does the case report provide takeaway lessons?** |
| --- | --- | --- | --- | --- | --- | --- | --- | --- |
| Du (2016) | Yes | Yes | Yes | Yes | Yes | Yes | Yes | Yes |
