## Supplementary material for "A systematic review of the experiences of minority language users in health and social care research": Table 1

### Table 1 List of papers by author, country, date and domain of interest

| **#** | **Author(s)** | **Country** | **Year** | **Domain of interest** | **Study participants** | **Study design** |
| --- | --- | --- | --- | --- | --- | --- |
| 1. | **Hunt and Bhopal**  (Hunt & Bhopal, 2004) | Scotland, UK | 2004 | Disparities in health care research. | This was a methodological commentary regarding including non-English speakers in research. | Research synthesis |
| 2. | **Gibbs et al**  (Gibbs, Nsiah-Jefferson, McHugh, Trivedi, & Prothrow-Stith, 2006) | USA | 2006 | Disparities in health care research. | N = 4 policies from the USA were analysed for this review. | Research synthesis |
| 3. | **Jacobs et al**  (Jacobs et al., 2006) | USA | 2006 | Disparities in health care research. | N = 2 systematic reviews of the literature. | Research synthesis |
| 4. | **Sue and Dhindsa**  (Sue & Dhindsa, 2006) | USA | 2006 | Disparities in health care research. | This was a review addressing ethnic and racial health and health care disparities research. | Research synthesis |
| 5. | **Beresford**  (Beresford, 2007) | England, UK | 2007 | Disparities in health care research. | This was a descriptive narrative to develop new directions in health inequalities research. | Research synthesis |
| 6. | **Lau et al**  (Lau, Chang, & Okazaki, 2010) | USA | 2010 | Disparities in health care research. | This was a commentary on randomised controlled trials looking at specific challenges facing investigators conducting ethnically inclusive trials. | Research synthesis |
| 7. | **Goode et al**  (Goode, Carter-Pokras, Horner-Johnson, & Yee, 2014) | USA | 2014 | Disparities in health care research. | This was a commentary on the lack of existing literature which focuses on the intersection of race, ethnicity, and disability. | Research synthesis |
| 8. | **Kaplan**  (Kaplan, 2014) | USA | 2014 | Disparities in health care research. | This was a commentary on the quality of data on “race” and “ethnicity” in the USA. | Research synthesis |
| 9. | **Coustasse et al**  (Coustasse, Bae, Arvidson, & Singh, 2010) | USA | 2010 | Disparities in health care use | N = 31,875 | Survey study |
| 10. | **Wilk et al**  (Wilk, Maltby, & Phillips, 2018) | Canada | 2018 | Disparities in health care use. | From the 2006 Aboriginal Peoples Surveys (APS) survey, 20,720 respondents were included and 24,150 from the 2012 APS. | Cross-sectional study |
| 11. | **Chirewa, 2012**  (Chirewa, 2012) | UK | 2012 | Disparities in health care use. | The research participants were drawn from the National Government Organisations (NGO’s) forum affiliates (n = 30), NHS primary care trusts (PCTs) (n = 2) and an expert group (academics) (N = 4). | Mixed method study |
| 12. | **Gaston-Johansson et al**  (Gaston-Johansson, Hill-Briggs, Oguntomilade, Bradley, & Mason, 2008) | USA | 2008 | Disparities in health care use. | N = 9 participants in each of the 6 focus groups. (N=42 in total) | Qualitative study |
| 13. | **Akhavan and Karlsen**  (Akhavan & Karlsen, 2013) | Sweden | 2013 | Disparities in health care use. | N = 5 x ‘migrant’ health service clients and 5 x physicians. | Qualitative study |
| 14. | **Hahm et al**  (Hahm, Lahiff, Barreto, & Chen, 2008) | USA | 2008 | Disparities in health care use. | N = 2,230 ethnic minority adolescents were interviewed over the telephone. | Survey study |
| 15. | **Di Pietro & Illes**  (Di Pietro & Illes, 2014) | Canada | 2014 | Disparities in health care use. | N = 52 reports published since 1981 were included in this systematic review. | Systematic review |
| 16. | **Strohschein et al**  (Strohschein, Merry, Thomas, & Gagnon, 2010) | Canada | 2010 | Maternal health | For the PACBIRTH study, 5 refugee/asylum seeking monolingual women per language, speaking Hindi, Tamil, Urdu, Spanish, and French participated in the testing. For the KAP study, 3 monolingual French-speaking and refugee/ asylum-seeking women participated, as well as 10 refugee/asylum-seeking monolingual Urdu, Tamil, and Hindi-speaking couples. Monolingual participants came from Mexico, Peru, Colombia, Pakistan, Sri Lanka, India, Cameroon, and the Congo | Qualitative study |
| 17. | **Johnsen et al**  (Johnsen et al., 2020) | Denmark | 2020 | Maternal health | N=18 Midwives | Qualitative study |
| 18. | **Huang et al**  (Huang et al., 2019) | China | 2019 | Maternal health | N = 10 qualitative papers were included in this systematic review | Systematic review |
| 19. | **Durbin et al**  (Durbin, Sirotich, & Durbin, 2017) | Canada | 2017 | Mental health | N=1449 mental health services clients. | Cross-sectional study |
| 20. | **Sadavoy et al**  (Sadavoy, Meier, & Ong, 2004) | Canada | 2004 | Mental health | N = 10 in Chinese speaking focus groups  N = 7 in Tamil speaking focus groups | Qualitative study |
| 21. | **Shattell et al**  (Shattell, Hamilton, Starr, Jenkins, & Hinderliter, 2008) | USA | 2008 | Mental health | N = 7 community members (2 x male and 5 x female)  N = 1 health educator  N = 1 doctoral student in Nursing  N = 2 undergraduate nursing students  N = 1 principal investigator | Qualitative study |
| 22. | **De La Torre**  (De La Torre, 2009) | USA | 2009 | Mental health | N = 20 Hispanic adults of both sexes (10 males and 10 females) | Qualitative study |
| 23. | **Rose and Cheung**  (Rose & Cheung, 2012) | USA | 2012 | Mental health | N = 54 articles published between 2001 and 2011. | Research synthesis |
| 24. | **Kim et al**  (Kim et al., 2011) | USA | 2011 | Mental Health | N=372 in total (Latino, Hispanic and Asian adults). | Survey study |
| 25. | **Brisset et al**  (Brisset et al., 2014) | Canada | 2014 | Mental health | N = 113 primary care practitioners in Montreal. | Survey study |
| 26. | **Lu et al**  (Lu, Dear, Johnston, Wootton, & Titov, 2014) | Australia | 2014 | Mental Health | N = 1449 mental health services clients | Survey study |
| 27. | **Woodall et al**  (Woodall, Morgan, Sloan, & Howard, 2010) | England, UK | 2010 | Mental health | N = 49 papers were included in this systematic review. | Systematic review |
| 28. | **Brown et al**  (Brown, Marshall, Bower, Woodham, & Waheed, 2014) | UK | 2014 | Mental health | N = 9 papers included in the systematic review. | Systematic review |
| 29. | **Tadić et al**  (Tadić et al., 2010) | UK | 2010 | Methodology in health research | N = 32 – interviewed (in stage 1)  N = 44 (in stage 2). | Mixed method study |
| 30. | **Mowlabaccus and Jodheea-Jutton**  (Mowlabaccus & Jodheea-Jutton, 2020) | Mauritius | 2020 | Methodology in health research | Qualitative: N = 23 open-ended question on-line survey.  Survey: N= 350 completed questionnaires | Mixed method study |
| 31. | **Dingoyan et al**  (Dingoyan, Schulz, & Mosko, 2012) | Germany | 2012 | Methodology in health research | The number of participants varied between 7 and 12 individuals per focus group. | Qualitative study |
| 32. | **Schildmann et al**  (Schildmann et al., 2016) | Germany and England | 2016 | Methodology in health research | N = 15 German and N =10 UK interviews were conducted. | Qualitative study |
| 33. | **Squires et al**  (Squires et al., 2019) | USA | 2019 | Methodology in health research | N = 35 home health care providers. | Qualitative study |
| 34. | **Murray and Buller**  (Murray & Buller, 2007) | England, UK | 2007 | Methodology in health research | N = 207 ‘original research pieces’ published in the BMJ were discussed in this review. | Research synthesis |
| 35. | **Wells and Zebrack**  (Wells & Zebrack, 2007) | USA | 2007 | Methodology in health research | In this review previous research was not selected but the authors utilized a social-ecological perspective to describe their findings. | Research synthesis |
| 36. | **Martinez**  (Martinez, Carter-Pokras, & Brown, 2009) | USA | 2009 | Methodology in health research | This was a review describing the lessons learned in using a participatory approach. | Research synthesis |
| 37. | **Yildiz and Bartlett**  (Yildiz & Bartlett, 2011) | England, UK | 2011 | Methodology in health research | N = 27 studies were included in this review. | Research synthesis |
| 38. | **Nishita and Browne**  (Nishita & Browne, 2013) | Hawaii, USA | 2013 | Methodology in health research | This was a broad literature review. | Research synthesis |
| 39. | **Waheed et al**  (Waheed, Hughes-Morley, Woodham, Allen, & Bower, 2015) | England, UK | 2015 | Methodology in health research | N = 9 studies were included. | Research synthesis |
| 40. | **Morville and Erlandsson**  (Morville & Erlandsson, 2016) | Sweden | 2016 | Methodology in health research | N = 21 articles were included in this literature review. | Research synthesis |
| 41. | **Premji et al**  (Premji, Kosny, Yanar, & Begum, 2020) | Canada | 2020 | Methodology in health research | N = 6 studies by the author were included in this review. | Research synthesis |
| 42. | **Mosconi**  (Mosconi et al., 2016) | Italy | 2016 | Methodology in health research | N = 1,852 survey responses. | Survey study |
| 43. | **MacFarlane et al** (MacFarlane, Singleton, & Green, 2009) | Ireland and England | 2009 | Migrant people’s health care needs. | Ireland: N=26 Serb Croat and Russian speaking refugees and asylum seekers (n = 16 females and n = 10 males).  England:  Focus groups (11 focus groups; n = 61 participants)  Semi-structured interviews (n = 28 participants). | Qualitative study |
| 44. | **Doyle et al**  (Doyle, Rager, Bates, & Cooper, 2013) | USA | 2013 | Migrant people’s health care needs. | N=9 Healthcare providers  N=11 Social service providers  N=20 migrant and seasonal farm workers  (N = 40 in total) | Qualitative study |
| 45. | **Betancourt et al** (Betancourt, Frounfelker, Mishra, Hussein, & Falzarano, 2015) | USA | 2015 | Migrant people’s health care needs. | Free List Group  N=39 = Somali Bantu  N=62 = Bhutanese refugees  Key Informant Group  N=21 = Somali Bantu  N=40 = Bhutanese refugees | Qualitative study |
| 46. | **Hunter-Adams and Rother**  (Hunter-Adams & Rother, 2017) | South Africa | 2017 | Migrant people’s health care needs. | Semi-structured interviews:  Congolese (n = 7)  Somali (n = 8) Zimbabwean (n = 8) women living in Cape Town  N = 9 Focus groups including men and women. | Qualitative study |
| 47. | **Chowdhury**  (Chowdhury, Naeem, Ferdous, Chowdhury, & Goopy, 2021) | Canada | 2021 | Migrant people’s health care needs. | N= 31 studies were included in this systematic review. | Systematic review |
| 48. | **Kale & Syed**  (Kale & Syed, 2010) | Norway | 2010 | Minority participation in health care | N = 453 participants from both primary and specialized healthcare facilities. | Cross-sectional study |
| 49. | **Angus et al**  (Angus et al., 2013) | Canada | 2013 | Minority participation in health care | N = 35 qualitative studies were included in this meta- analysis. | Meta analysis |
| 50. | **Wang and Kwak**  L. Wang & Kwak, 2015) | Canada | 2015 | Minority participation in health care | N = 8 focus groups (n=54 in total; 81.5% female and 18.5% male). | Mixed method study |
| 51. | **Wang**  (A. Wang et al., 2019) | Canada | 2019 | Minority participation in health care | Quantitative chart review: n = 2420 registered charts  Qualitative focus groups: n = 13 participants | Mixed method study |
| 52. | **Claydon-Platt et al** (Claydon-Platt, Manias, & Dunning, 2013) | Australia | 2013 | Minority participation in health care | N = 11 people with diabetes, N = 10 carers and N = 10 health professionals were interviewed. (N= 31 in total) | Qualitative study |
| 53. | **Tatari et al**  (Tatari et al., 2020) | Denmark | 2020 | Minority participation in health care | N = 37 women from ten different non-Western countries participated in the study. | Qualitative study |
| 54. | **Castillo**  (Castillo, Gandy, Bradko, & Castillo, 2019) | USA | 2019 | Minority participation in health care | N = 18 papers included in the systematic review. | Systematic review |
| 55. | **Blanchet et al**  (Blanchet et al., 2017) | Canada | 2017 | Minority participation in health care research | N = 259 Parent and child dyads (251 biological mothers, one adoptive mother, three fathers). | Cohort Study |
| 56. | **Eriksson-** **Sjöö et al**  (Eriksson-Sjöö, Cederberg, Östman, & Ekblad, 2012) | Sweden | 2012 | Minority participation in health care research | N = 78 newly‐arrived Arabic‐speaking adult refugees in Malmö, Sweden. | Mixed method study |
| 57. | **Ahlmark et al**  (Ahlmark, Algren, Holmberg, & Norredam, 2014) | Denmark | 2014 | Minority participation in health care research | N = 177,639 (survey).  Also data from 10 immigrants and 13 descendants between the ages of 18 to 54. | Mixed method study |
| 58. | **Robinson and Trochim**  (Robinson & Trochim, 2007) | USA | 2007 | Minority participation in health care research | N=20 steering committee members (n =20),  N=16 community advisory board members regional N N = 6 advisory board members  N = 5 lay community members (n=5) | Qualitative study |
| 59. | **Fisher**  (Fisher, 2011) | Australia | 2011 | Minority participation in health care research | N=54 from five communities in Australia.  N = 24 from health and support agency staff who provide services to them.  Agency support staff represented a range of professional perspectives.  (N=78 in total) | Qualitative study |
| 60. | **French and Stavropoulou**  (French & Stavropoulou, 2016) | UK | 2016 | Minority participation in health care research | N = 12 specialist nurses representing 7 different clinical specialties and 7 different NHS Trusts. | Qualitative study |
| 61. | **O’Connor et al**  (O’Connor, Adem, & Starks, 2018) | USA | 2018 | Minority participation in health care research | Community leader interviews (n = 6) and focus groups with lay members (n = 16) from the three largest East African communities in the Seattle area (Eritrean, Ethiopian and Somali) | Qualitative study |
| 62. | **Greene et al**  (Greene, Karavatas, Cooper, & Zamorano-Torres, 2013) | USA | 2013 | Minority participation in health care research | N = 30 patients from the Washington, DC metropolitan area, whose primary language is Spanish | Survey study |
| 63. | **Carlini et al**  (Carlini, Safioti, Rue, & Miles, 2015) | USA | 2015 | Minority participation in health care research | Brazilian community members living in the USA: (Florida, California and New Jersey). | Survey study |
| 64. | **Falla et al**  (Falla, Veldhuijzen, Ahmad, Levi, & Richardus, 2017) | The Netherlands | 2017 | Minority participation in health care research | N = 238 respondents to on-line survey. | Survey study |
| 65. | **Du**  (Du, 2018) | USA | 2018 | Providing health care services to ethnic minority patients. | N = 1. This case report was a reflection of a graduate medical student who provided real-life examples regarding language and communication. | Case report |
| 66. | **Peek et al**  (Peek et al., 2012) | USA | 2012 | Providing health care services to ethnic minority patients. | N = 167 physician organisations (of differing levels of membership). | Cross-sectional study |
| 67. | **Silveira et al**  (Silveira et al., 2020) | USA | 2020 | Providing health care services to ethnic minority patients. | N =-16,415 Hispanic/Latino adults in the U.S. | Cross-sectional study |
| 68. | **Bhuiyan et al**  (Bhuiyan, Urmi, Chowdhury, & Rahman, 2019) | Bangladesh | 2019 | Providing health care services to ethnic minority patients. | N = 50 participants of different age groups.  N= 44 male  N = 6 female | Mixed method study |
| 69. | **Tan and Denson**  (Tan & Denson, 2019) | Australia | 2019 | Providing health care services to ethnic minority patients. | N = 38 bilingual/multilingual psychologists working in Australia in 2015.  N = 11 participants undertook supplementary telephone interviews | Mixed method study |
| 70. | **Vandan et al**  (Vandan et al., 2020) | Hong Kong, China | 2020 | Providing health care services to ethnic minority patients. | N = 22 Health care professionals | Qualitative study |
| 71. | **Schwei et al**  (Schwei, Del Pozo, Agger-Gupta, Alvarado-Little, Bagchi, Hm Chen, et al., 2016) | USA and Canada | 2016 | Providing health care services to ethnic minority patients. | N = 136 studies prior to 2003 and N = 426 studies from 2003 to 2010. | Research synthesis |
| 72. | **Joo and Liu**  (Joo & Liu, 2020) | South Korea | 2020 | Providing health care services to ethnic minority patients. | N = 8 papers were included in this systematic review. | Systematic review |
| 73. | **Clarke et al**  (Clarke et al., 2013) | USA | 2013 | Racial and ethnic gaps in health care. | N = 11 systematic reviews were included in this meta analysis. | Meta analysis |
| 74. | **Haley et al**  (Haley, Southwick, Parikh, Farrar-edwards, & Boden-albala, 2017) | USA | 2017 | Racial and ethnic gaps in health research. | N = 29 clinical research coordinators (CRCs) | Qualitative study |
