## Supplementary material for "A systematic review of the experiences of minority language users in health and social care research": Table 2

**Table 2	Themes and sub-themes for minority language speakers in health research**

| **Theme** | **Sub-theme** | **Number of papers** |
| --- | --- | --- |
| 1. Disparities in health care | Disparities in health care research | 8 |
|  | Disparities in health care use | 7 |
| 2. Maternal health |  | 3 |
| 3. Mental health |  | 10 |
| 4. Methodology in health research |  | 14 |
| 5. Migrant and minorities in health care | Migrant people’s health care needs | 5 |
|  | Minority participation in health care | 7 |
|  | Minority participation in health care research | 10 |
|  | Providing health care services to ethnic minority patients | 8 |
| 6. Racial and ethnic gaps | Racial and ethnic gaps in health care | 1 |
|  | Racial and ethnic gaps in health research | 1 |
|  | **Total** | **74** |
